# Multi-Scale Characterization of Distance-dependent Functional Dysconnections in Major Depressive Disorder Across Two Asian Cohorts: A Genomic, Neurochemical, and Cellular Perspective

**DOI:** 10.1101/2024.11.20.24317665

**Authors:** Rui Qian, Huaijin Gao, Bowen Qiu, Zichao Li, Baorong Gu, Tongmei Ye, the DIRECT Consortium, Dan Wu, Zhiyong Zhao

**Author notes:** Corresponding to: Zhiyong Zhao, PhD.

## Abstract

Major Depressive Disorder (MDD) is a prevalent, chronic, and multidimensional mental disorder characterized by widespread functional dysconnectivity in the whole brain. However, the potential molecular, cellular, and neural mechanisms, contributing to the diverse symptomatology and heterogeneity of MDD remain poorly understood. This study aims to elucidate the multi-scale pathophysiological mechanisms underlying MDD subtypes by integrating functional connectivity, transcriptomic, neurotransmitter, and cell-type analyses across two Asian cohorts: the Chinese REST-meta-MDD Consortium (Discovery) and the Japanese Decoded Neurofeedback Project (Validation). The discovery cohort identified distinct patterns of distance-dependent functional connectivity strength (FCS) alterations in MDD, revealing short- to medium-range hyperconnectivity in both total MDD and recurrent MDD (RMDD) patients, with long-range hyperconnectivity specifically observed in RMDD. In contrast, first-episode drug-naïve (FEDN) patients did not exhibit significant distance-dependent alterations in FCS. Genes associated with the FCS differences between FEDN and RMDD were enriched in pathways related to chemical synaptic transmission, neuron projection, and synaptic signaling. Moreover, FCS alterations in MDD subtypes were correlated with neurotransmitter receptor densities, particularly in the monoaminergic (e.g., 5HT1a, 5HT2a, and KappaOp) and GABAergic (GABAa) systems. Distinctive cell-type associations were observed, with astrocytes, endothelial cells, and oligodendrocyte precursor cells (OPCs) linked to FCS changes in RMDD, while only OPCs were associated with alterations in FEDN. The validation cohort partially replicated the key findings regarding distance-dependent FCS alterations, transcriptomic signatures, neurotransmitter associations, and cell-type specific relationships. These findings provide novel insights into the neurobiological underpinnings of functional dysconnections in MDD subtypes.

## 1. INTRODUCTION

Major depressive disorder (MDD) is among the leading causes of disease burden worldwide, and it will rank first by 2030 according to the World Health Organization (WHO) ^1^. Compared to first-episode drug-naïve MDD (FEDN), patients with recurrent MDD (RMDD) experience more severe cognitive impairments and an increased likelihood of suicidal thoughts ^2,3^. However, the neurobiological mechanisms that differentiate these MDD subtypes remain poorly understood. Recent studies have highlighted different functional connectivity (FC) changes in RMDD and FEDN patients, with RMDD exhibiting decreased FC in regions where FEDN shows increased FC, particularly within the default mode network, indicating distinct and complex mechanisms underlying these disease subtypes ^4,5^. These abnormal connections are likely influenced by underlying molecular and cellular processes, which in turn shape the brain’s functional architecture ^6–8^. Characterizing the molecular and cellular substrates of brain dysconnectivity across subtypes is crucial for developing biomarkers and identifying novel therapies for MDD ^9,10^.

Functional connectivity strength (FCS) is a widely used fMRI metric that quantifies the functional importance and information processing load of each brain region with the other regions in whole brain ^11^. Previous studies have consistently reported FCS alterations in MDD patients compared to healthy controls across multiple brain networks and specific regions implicated in emotional, cognitive, and functional disturbances associated with depression ^12–14^. However, most prior studies have treated FCS as a spatially homogeneous metric, neglecting its inherently distance-dependent properties. Emerging evidence suggest that short-range connections primarily manifest in emotion-regulation areas, such as the limbic system and default mode network, while long-range connections impact broader cognitive and attention networks, particularly in MDD ^15–17^. This distance-dependent approach has provided novel insights into the neural mechanisms underlying various psychiatric disorders, including schizophrenia (SCZ), bipolar disorder (BD), and MDD ^16^. However, it remains unclear whether distance-dependent FCS alterations exhibit different patterns in the MDD subtypes of FEDN and RMDD, although they have been documented in both drug-naïve and treated MDD patients ^18,19^.

The Allen Human Brain Atlas (AHBA) has revolutionized our ability to link gene expression patterns with neuroimaging phenotypes ^20–22^, as cortical gene expression patterns are highly conserved across individuals ^23^. Using transcription-neuroimaging methods, studies have identified significant associations between gene expression and brain characteristics such as cortical volume, structural connectivity, and functional connectivity. These studies revealed higher transcriptomic profile similarity within functional networks and specific gene sets related to corticocortical network architecture ^24–26^. Crucially, a recent study has identified biological and cellular pathways consistent with the known biology of a wide range of molecular neuroimaging markers through transcriptomic decoding of the regional distribution of well-known molecular markers ^27^, further validating the transcription-neuroimaging association approach. In the context of MDD, recent studies have linked gene expression profiles to neuroimaging alterations, including cortical thinning, morphometric similarity networks, and functional connectivity changes ^28,29^. Although studies have explored the molecular basis of distinct neuroimaging changes in first-episode MDD compared to healthy controls ^30^, the transcriptional mechanisms underlying MDD subtypes (i.e., FEDN vs. RMDD) remain unclear.

Neuroimaging abnormalities in MDD may reflect disrupted neurotransmitter systems orchestrating neural signaling across brain circuits ^31–33^. Molecular imaging studies have revealed serotonergic, dopaminergic, glutamatergic, and GABAergic dysregulations in MDD pathophysiology, including reduced serotonin transporter binding in the amygdala and midbrain, and altered glutamate levels in the prefrontal and anterior cingulate cortices ^34,35^. Recent advances in computational biology have enabled estimation of cell-type-specific gene expression patterns in the human brain, illuminating the contributions of neurons, glia, and other cell types to MDD pathophysiology ^36,37^. Postmortem studies have identified alterations in astrocyte and microglial markers in MDD patients, suggesting a critical role for glial cells in this disorder ^38^. However, the relationships between aberrant FCS in MDD subtypes and neurotransmission, as well as cell types, remain elusive. Investigating these intricate associations could provide vital insights into the cellular and molecular underpinnings of brain dysfunction in MDD.

Variations in demographic factors significantly impact the manifestation and etiology of MDD through complex interactions among sociopsychological factors, lifestyle patterns, and genetic predispositions ^39–41^. Current large-sample neuroimaging studies in MDD and its subtypes have primarily focused on Western populations, with relatively few studies involving Asian populations, despite evidence of differences between them in brain function and structure ^42,43^. Existing large-sample databases such as the UK Biobank ^44^, the ENIGMA MDD Working Group ^43,45^, and the PsyMRI Consortium ^46^ have identified key structural and functional abnormalities in MDD patients, including cortical thinning, hippocampal volume reductions, and altered network connectivity. To expand this scope, our study utilized two major Asian databases: the Chinese REST-meta-MDD Consortium (Discovery) and the Japanese Decoded Neurofeedback (DecNef) Project (Validation), to investigate the neurobiological basis underlying MDD subtypes in Asian populations. We first explored distance-dependent FCS alterations in the discovery cohort, which included FEDN and RMDD subtypes. Then, by integrating multivariate partial least squares regression, cross-modal spatial correlation, and cell-type-specific analyses, we delineated the genomic, neurochemical, and cellular underpinnings of functional dysconnectivities induced by MDD subtypes. Finally, we replicated all analyses in the validation dataset to verify the findings observed in the discovery cohort. An overview of the data processing is presented in Fig. 1. This multi-scale approach bridges macroscale-level connectional alterations and microscale-level molecular and cellular features, offering valuable insights into the pathophysiological mechanisms of MDD in Asian populations and potentially guiding future therapeutic strategies.

**Fig. 1.**
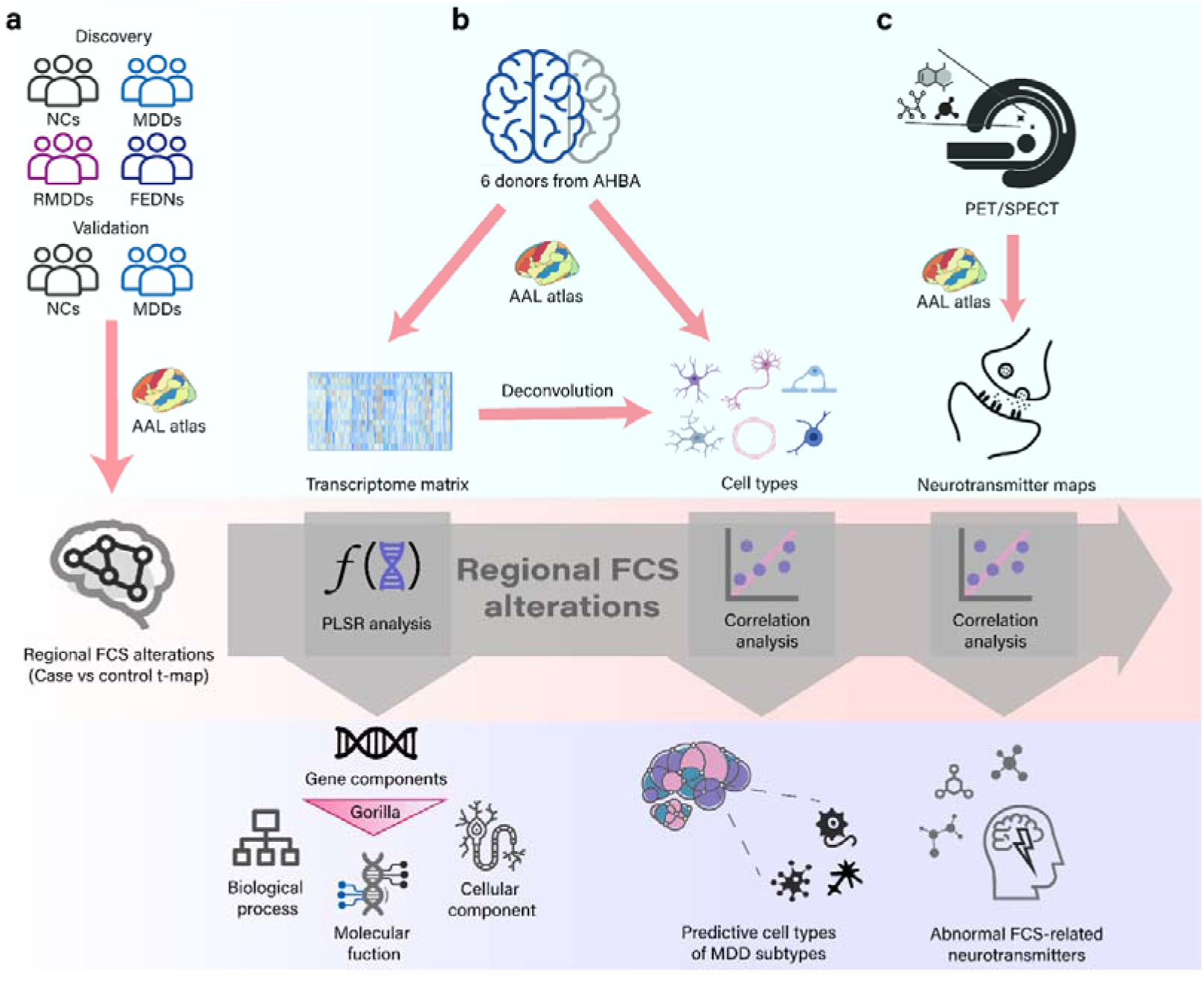
Study Overview. **a** Regional functional connectivity strength (FCS) analysis. Functional MRI data from discovery and validation cohorts, including overall MDD, FEDN, RMDD, and normal controls (NCs). Individual FCS was constructed based on a 116 ×116 functional connectivity matrix and produced a 116 ×N (where 116 represents the number of regions of interest from the AAL atlas and N is the number of subjects) FCS matrix. Then, FCS differences between groups were calculated using a linear mixed model. **b** Gene expression profiles and cell-type analysis. Transcriptome profiles from the Allen Human Brain Atlas in 116 regions (left hemisphere only) were averaged across six postmortem brains. We employed partial least squares (PLS) regression to identify imaging-transcriptomic associations and conducted enrichment analysis on the gene list associated with the first component of PLS (PLS1). The distribution of six cell types was obtained through deconvolution from the transcriptome matrix, followed by correlation analysis between cell-type distributions and FCS alterations. **c** Neurotransmitter density profiles. Correlation analysis assessed the associations between functional dysconnectivity and neurotransmitter maps from PET/SPECT.

## 2. MATERIALS AND METHODS

### 2.1 Subject

Resting-state fMRI data from the REST-meta-MDD Consortium ^47^, which includes 1,300 patients MDD and 1,128 controls from 25 sites, were screened to form the discovery cohort. The exclusion criteria were as follows: (1) sites with late-onset depression or remitted patients; (2) subjects with missing demographic information (sex, age, and education) or poor-quality imaging; (3) subjects younger than 18 or older than 65 years; (4) subjects with inadequate brain coverage, excessive head motion, or poor spatial correlation with the group mean ReHo map; and (5) sites with fewer than 10 subjects per group. Ultimately, the discovery cohort comprised 848 MDD patients and 794 normal controls (NCs) from 17 sites. Four paired groups were established: 232 FEDN patients and 394 NCs, 227 FEDN patients and 100 first-episode medicated (FEM) patients, 189 recurrent MDD (RMDD) patients and 427 NCs, and 119 FEDN patients and 72 RMDD patients. All sites provided data on diagnosis, age at scan, sex, and education. All study participants gave written informed consent at their local institutions.

The validation cohort consisted of inter-regional correlation matrices derived from resting-state fMRI data collected from the DecNef Project at the University of Tokyo Hospital between 2014 and 2017. The matrices were calculated for seven diagnostic categories (normal, autism spectrum disorder, major depressive disorder, schizophrenia, bipolar disorder, dysthymia, and other psychiatric disorders) using three anatomical atlases (AAL, R93, and BAL). Due to the high missing rate in transcriptome data for the other two atlases, only the AAL atlas was used in this study. We included MDD patients and NCs aged 18 to 65 years, with no contraindications for MRI and no excessive head motion (frame-wise displacement > 0.5 mm) during fMRI scans. The final sample consisted of 62 MDD patients and 170 NCs, matched for age, sex, and handedness, from a single site. Note that the MDD patients did not have information of FEDN and RMDD in the validation cohort.

### 2.2 MRI Data Acquisition and Processing

Details of MRI data acquisition are presented in the Supplementary Material. The fMRI data across all sites in the discovery cohort were preprocessed consistently ^47^ as follows: (i) The first 10 volumes were discarded, and slice-timing correction was applied. (ii) Images for each subject underwent realignment using a six-parameter linear transformation. (iii) T1-weighted images were co-registered to the mean functional image without resampling and were then segmented into gray matter (GM), white matter (WM), and cerebrospinal fluid (CSF). (iv) Functional images were transformed from individual native space to Montreal Neurological Institute (MNI) space using the DARTEL method. (v) Confounding factors, including head motion, WM, and CSF signals, were removed. (vi) Spatial smoothing was applied using an isotropic Gaussian kernel of 6 mm full width at half maximum (FWHM), and temporal bandpass filtering (0.01–0.1 Hz) was performed on all time series.

The validation data were preprocessed using a similar pipeline ^48^ with the one used in the discovery data, including excluding the first 10 volumes, slice-timing correction, realignment, co-registration, distortion correction, segmentation of T1-weighted images, normalization to MNI space, spatial smoothing with a 6 mm isotropic Gaussian kernel, and bandpass filtering between 0.01 and 0.08 Hz.

### 2.3 Distance-dependent FCS Calculation

Regions of interest (ROIs) were defined using the AAL atlas ^48^. First, we obtained the functional connectivity (FC) matrix by calculating Pearson’s correlation coefficients between the time series of fMRI signals for paired ROIs. After computing the FC matrix, we took the absolute values of the correlation coefficients. Connections above a threshold of 0.25 were set to 1, while those below were set to 0 to exclude spurious connections ^49^. The FCS value for a single ROI is defined as the sum of its direct connections to other ROIs. In an undirected network, the FCS value of one node (ROI) can be measured as follows:

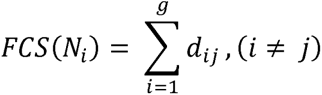

where *FCS*(*N_i_*) represents the FCS value of node *N_i_*, *g* indicates the total number of nodes in the graph network, and *d_ij_* denotes the number of edges (connections) between *N_i_* and the other *g*-1 nodes, with *i* ≠*j* to exclude self-connections. This approach allowed us to generate individual-level FCS maps, which were subsequently normalized to Z-score maps by subtracting the mean FCS value across all nodes and dividing by the standard deviation of FCS values.

To compute distance-dependent FCS, the three-dimensional anatomical distance between every pair of voxels (*i* and *j*) was approximated using the Euclidean distance:

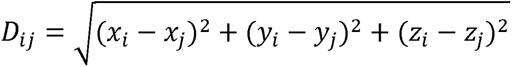

The stereotaxic coordinates for voxels *i* and *j* in MNI space are represented as (x*_i_*, y*_i_*, z*_i_*) and (x*_j_*, y*_j_*, z*_j_*), respectively. Whole-brain functional connectivity was divided into 13 bins, with Euclidean distances categorized into 10-mm increments ranging from 0 to 130 mm ^16^. We calculated the FCS matrix for each bin using the same approach as for the whole-brain FCS, resulting in a distance-dependent FCS matrix for each subject.

### 2.4 Gene Data, Neurotransmitters, and Cell Types

The human microarray-based gene expression data were downloaded from the AHBA^22^ (http://human.brain-map.org). The dataset includes tissue samples collected from the brains of six adult donors (mean age: 42.5 years; one female). We utilized the Abagen toolbox (https://www.github.com/netneurolab/abagen) to process and map the transcriptomic data onto 116 parcellated brain regions defined by the AAL atlas. Briefly, the preprocessing of gene expression data included the following steps ^30^: (i) updating probe-to-gene annotations; (ii) applying an intensity-based filter; (iii) selecting probes; (iv) matching samples to regions; (v) handling missing data; (vi) normalizing samples; (vii) normalizing genes; (viii) combining samples into region metrics; and (ix) selecting stable genes. Finally, we obtained a gene expression matrix of 116 regions × 15,633 genes for further analysis.

Neurotransmitter receptor and transporter density were obtained from a recent report ^50^ that collated data from a large number of PET studies involving over 1,200 healthy adults (42% female; mean sample-size weighted age = 36.6 years). This dataset provided the densities of nine neurotransmitter receptors and transporters across the whole brain, including dopamine, serotonin, glutamate, GABA, acetylcholine, opioids, cannabinoids, noradrenaline, and fluorodopa.

To estimate the densities of canonical cell types across brain regions, we applied a feature gene decomposition-based deconvolution method ^51^. This approach leverages known genetic markers to quantify the proportions of six principal cell types: neurons, astrocytes, oligodendrocytes, microglia, endothelial cells, and oligodendrocyte precursor cells (OPCs) ^37,52^. Using transcriptomic data from the AHBA, we calculated the densities of these six cell types across the 116 brain regions defined by the AAL atlas. The deconvolution was performed using the BRETIGEA software package (https://github.com/andymckenzie/BRETIGEA) ^37^.

### 2.5 Statistical Analysis

We compared FCS differences among five groups (total MDD vs. NC, FEDN vs. NC, FEM vs. FEDN, RMDD vs. NC, FEDN vs. RMDD) in the discovery cohort using a linear mixed model, controlling for age, sex, education, and head motion as covariates, and site as a random factor ^53^. For the validation cohort, we conducted two-sample t-tests (MDD vs. NC), controlling for age, sex, and education. All results were corrected by False Discovery Rate (FDR, p < 0.05) correction to control false positive discoveries.

A partial least squares regression (PLSR) analysis was performed to explore the association between transcriptional profiles and FCS differences. We first aligned the gene expression data (15,633 genes) with t-values of FCS from group comparisons ^54^, treating gene expression as predictor variables and FCS t-values as the response variable. Then we adopted a spatial autocorrelation-corrected permutation test to determine if the *R*^2^ of the PLSR component significantly exceeded chance levels^55^. For each significant component, we applied a bootstrapping method to correct for estimation errors in the weights of each gene ^56^. Finally, the genes were ranked based on corrected weights reflecting their contributions to the PLSR component, and enrichment analysis was conducted to identify enriched Gene Ontology terms using the Gorilla tool ^57^ (http://cbl-gorilla.cs.technion.ac.il/), considering all three ontology categories: biological process, molecular function, and cellular component.

We used the JuSpace toolbox ^58^ to examine spatial correlations between the t-values of group comparisons and various neurotransmitter systems. Specifically, we calculated Pearson correlation coefficients between the t-values and neurotransmitter densities across 116 brain regions. Statistical significance was assessed using exact p-values derived from spatial permutation-based null maps with 5,000 permutations, with a significance threshold set at p < 0.05. Additionally, we performed pairwise Pearson correlation analyses between the densities of the six cell types and the t-values of FCS, adjusting for multiple comparisons using FDR correction, with a significance threshold of 0.05.

## 3. RESULTS

### 3.1 Sample Composition

For the discovery cohort, total MDD patients and NCs were matched in age (MDD: 34.3 ± 11.5 years; NC: 34.4 ± 13.0 years; p = 0.313). However, MDD patients exhibited significantly fewer years of education compared to NCs (12.0 ± 3.4 vs. 13.6 ± 3.4 years; p < 0.001) and a higher proportion of females (63.5% vs. 57.9%; χ² = 7.945, p = 0.005). Both FEDN and RMDD patients had fewer years of education than NCs (FEDN vs. NC: 12.2 ± 3.4 vs. 13.6 ± 3.6 years, p < 0.001; RMDD vs. NC: 11.7 ± 3.2 vs. 13.4 ± 3.8 years, p < 0.001). FEDN patients were younger (35.4 ± 11.3 years) than RMDD patients (36.3 ± 12.7 years) and exhibited a shorter duration of illness (27.0 ± 39.5 vs. 88.7 ± 80.1 months; p < 0.001).

The validation cohort comprised 62 MDD patients (age range: 18-63 years) and 170 NC participants (age range: 19-80 years), with MDD patients being significantly older than NCs (38.74 ± 11.62 years vs. 35.59 ± 17.5 years; p < 0.001). The gender distribution indicated a higher proportion of males in the MDD group (58.06%) compared to the NC group (45.88%).

### 3.2 Distance-dependent FCS Differences Between MDD and NC Groups

Compared to NCs, increased FCS was observed in the left angular gyrus in the total MDD group and in the right posterior cingulate gyrus in the RMDD group (Fig. 2a). No significant FCS differences were observed in other subgroup comparisons. However, when accounting for the effect of spatial distance between regions on FC, we identified more MDD-related alterations and revealed three distinct patterns of distance-dependent FCS alterations in MDD (Fig. 2b). Specifically, compared to NCs, total MDD patients exhibited enhanced short-range (distance = 20-30 mm) FCS in wide brain regions, including the right precentral gyrus, bilateral superior frontal gyrus, right middle frontal gyrus, right supplementary motor area, left calcarine fissure and adjacent cortex, left cuneus, left superior parietal gyrus, bilateral paracentral lobule, and left superior temporal gyrus. In RMDD, short-range (distance = 30-40 mm) FCS was increased in the left inferior parietal lobe, left supramarginal gyrus, right inferior occipital gyrus, left superior occipital gyrus, and cuneus, while it was decreased in the left inferior temporal gyrus and bilateral frontal pole. Regarding medium-range (distance = 60-70 mm) connectivity, both total MDD and RMDD exhibited increased FCS in the left precentral gyrus and right inferior frontal gyrus, along with decreased FCS in the bilateral superior temporal pole. Additionally, RMDD also demonstrated increased FCS in the right superior parietal gyrus. Moreover, RMDD patients showed altered long-range (distance = 120-130 mm) FCS in several brain regions, with increased FCS in the left precentral gyrus, right inferior parietal lobe, bilateral inferior frontal gyrus, posterior cingulate gyrus, medial temporal lobe, right superior temporal gyrus, and decreased FCS in the right rectus gyrus and right middle temporal pole. No significant distance-dependent FCS differences were observed between FEDN and NC, between FEDN and FEM, and between FEDN and RMDD (Supplementary Fig 1).

**Fig.2.**
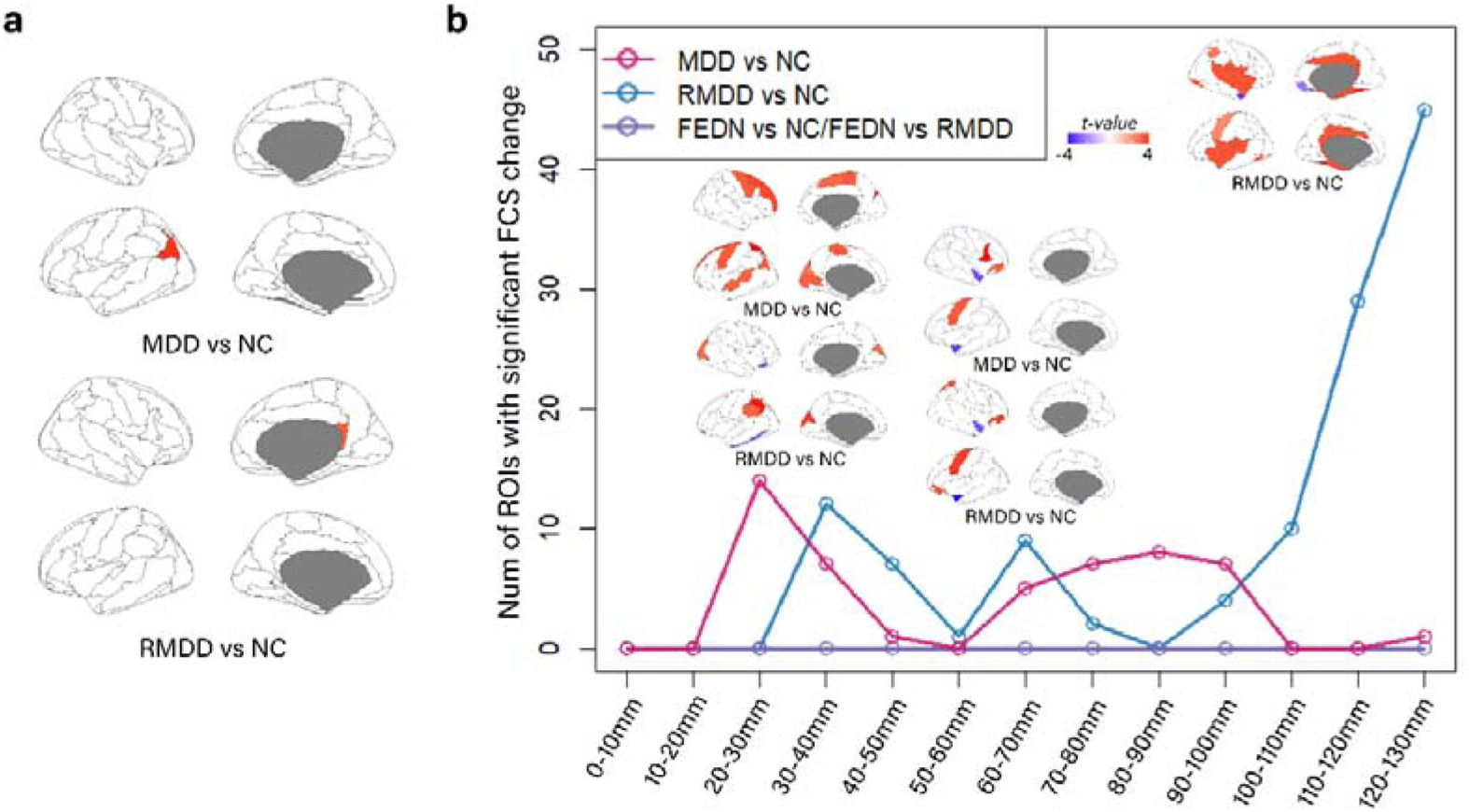
Distance-dependent FCS changes in the MDDs from discovery cohort. **a** Brain regions exhibiting significant FCS alterations without considering distance dependency in total MDD and RMDD compared to NC. **b** Distance-dependent FCS alterations demonstrating short, medium, and long-range heterogeneity in total MDD vs. NC (pink), FEDN vs. RMDD/FEDN vs. NC (purple), and RMDD vs. NC (blue). The x-axis represents the Euclidean distance between brain regions, ranging from 0 to 130 mm. The y-axis indicates the number of ROIs with significant FCS changes. Brain images illustrate areas showing significant differences between groups in FCS after FDR correction. The colorbar represents t values, with positive and negative values indicating higher and lower FCS in the former group relative to the latter group, respectively.

### 3.3 Association Between FCS Abnormalities in MDD Subtypes and Transcriptomic Signatures

The first component of PLSR (PLS1) has the highest explained variance (36.3%, p < 0.0001) for FEDN vs. RMDD. The regional mapping of these two components was positively correlated with the Z-map of the FCS difference (r = 0.48, p = 0.0001, permutation tests with spatial autocorrelation correction, Fig. 3a). FCS differences between groups showed positive correlations with expression values of some genes, such as CMTM4, PDE8B, and TMEM121, and negative correlations with expression values of other genes, including USP3, CECR2, and DPF3 (Fig. 3b). Additionally, Gene Ontology enrichment analysis revealed that the PLS1 genes were enriched in biological processes related to chemical synaptic transmission, as well as in molecular functions associated with chemical synaptic transmission and cell components of plasma membrane-bounded neuron projections (Fig. 3c). Furthermore, the PLS1 genes for FEDN vs. NC were enriched in biological processes related to the electron transport chain, molecular functions pertaining to NADH dehydrogenase activity, and cell components of mitochondrial inner membrane protein complexes and respiratory chain complexes (Supplementary Fig. 3 and Fig. 4). In contrast, genes for RMDD vs. NC were enriched in biological processes related to chemical synaptic transmission, molecular functions involving RNA polymerase II proximal promoter sequence-specific DNA binding, and cell components of the nucleus (Supplementary Fig. 3 and Fig. 5). Gene Ontology results for total MDD vs. NC and FEM vs. FEDN are presented in Supplementary Fig. 6 and Fig. 7, respectively.

**Fig.3.**
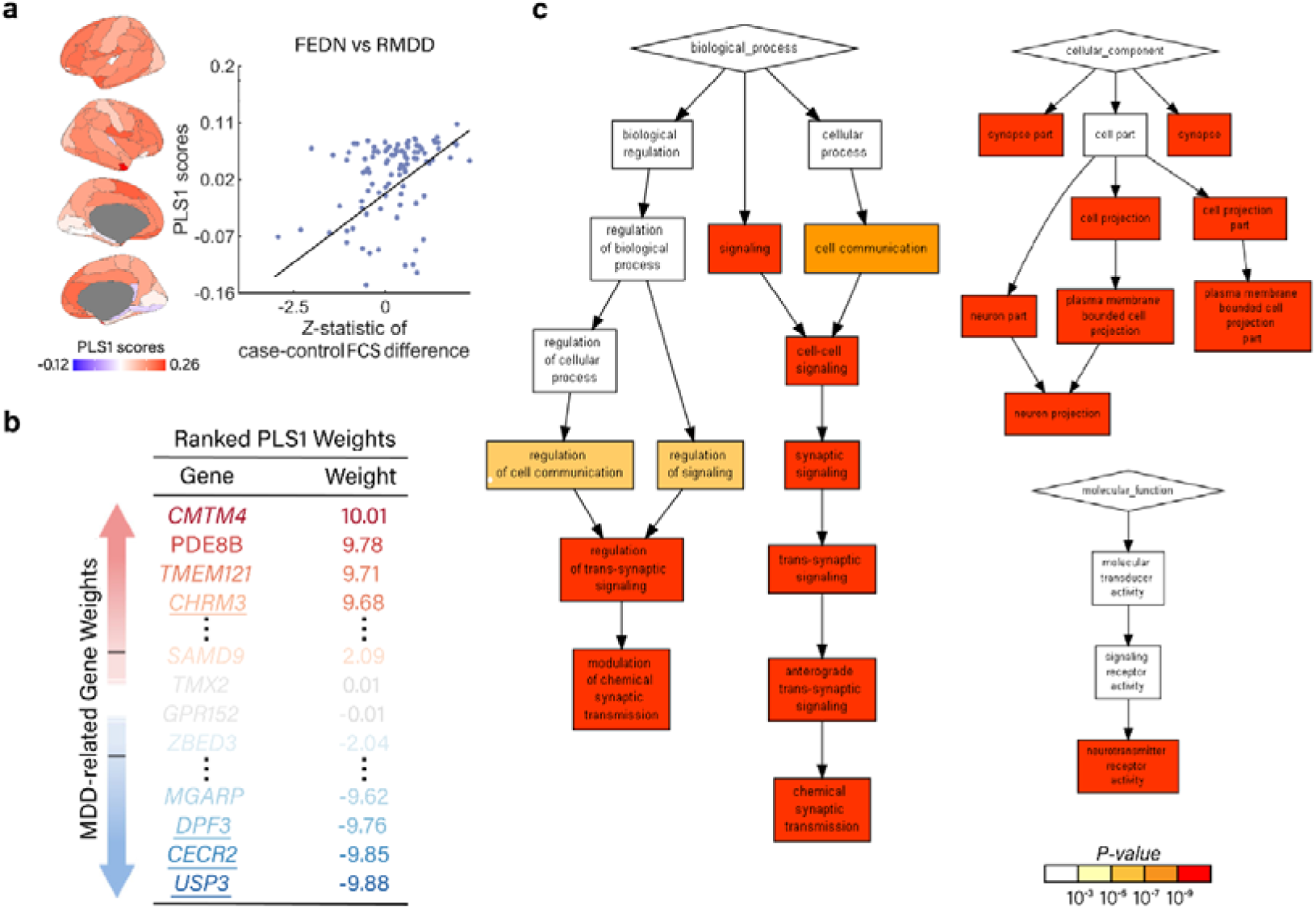
Association between FCS differences in FEDN vs. RMDD and transcriptomic signatures. **a** The first PLS component (PLS1) identified a gene expression profile exhibiting high expression across the cortex. The transcriptional profiles were positively correlated with the FCS differences between groups (permutation tests corrected 10,000 times). Each dot represents a brain region. **b** Ranked PLS1 loadings. **c** PLS1 gene enrichment analysis results via Gorilla (FDR correction, p < 0.05). PLS1 genes for FEDN vs. RMDD were enriched in biological processes associated with chemical synaptic transmission, molecular functions of chemical synaptic transmission, and cell component of plasma membrane-bounded neuron projections.

**Fig. 4.**
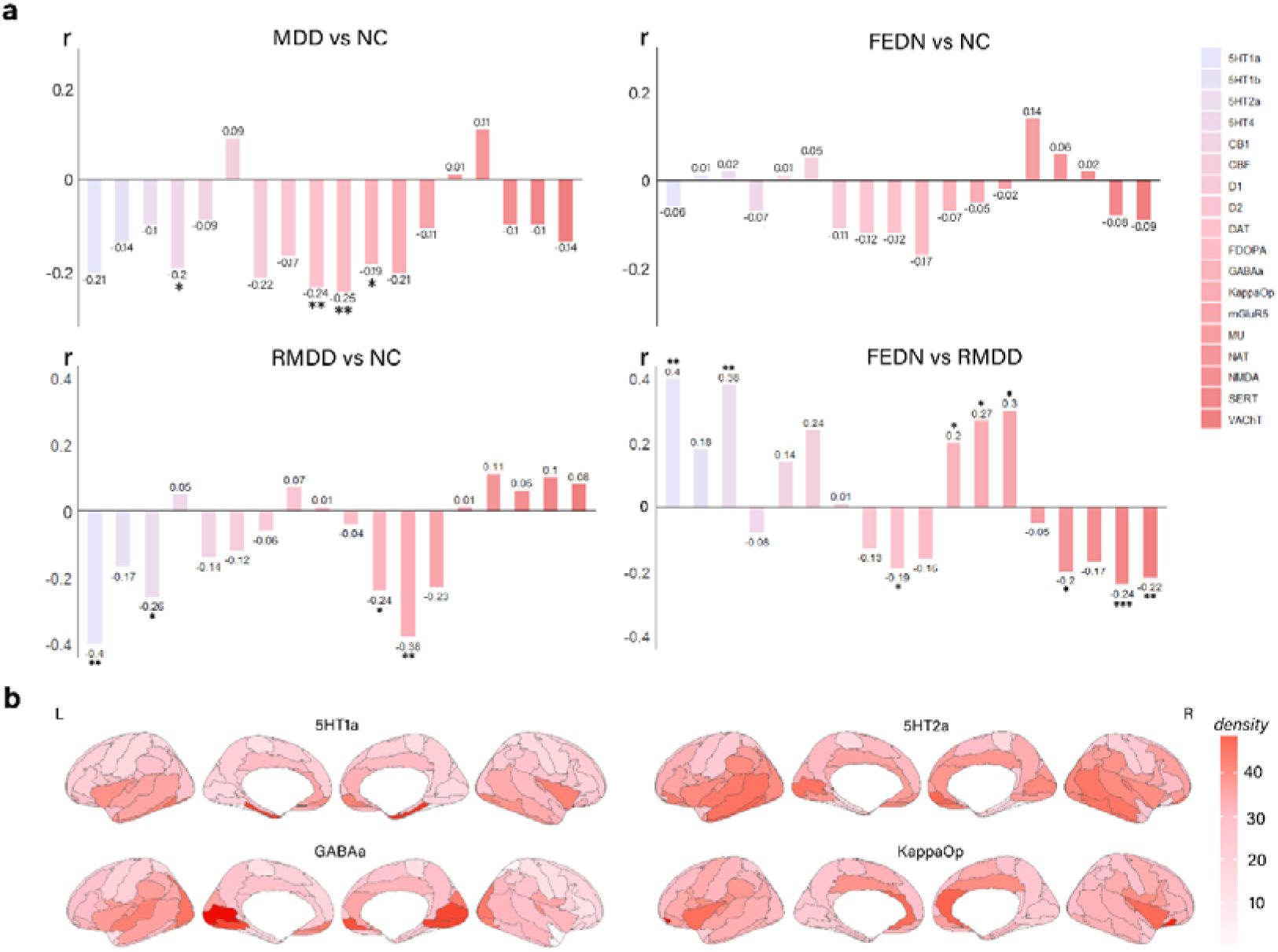
Associations between FCS alterations and the densities of neurotransmitter receptors/transporters. **a** Correlation trends between neurotransmitter densities and FCS differences in MDD vs. NC, FEDN vs. NC, RMDD vs. NC, and FEDN vs. RMDD groups. **b** Density distribution of 5-HT1a, 5-HT2a, GABA-a, and KappaOp, showing the most significant correlations with FCS differences between MDD subtypes. *: 0.01 < p < 0.05; **: 0.001 < p < 0.01; ***: p < 0.001.

**Fig. 5.**
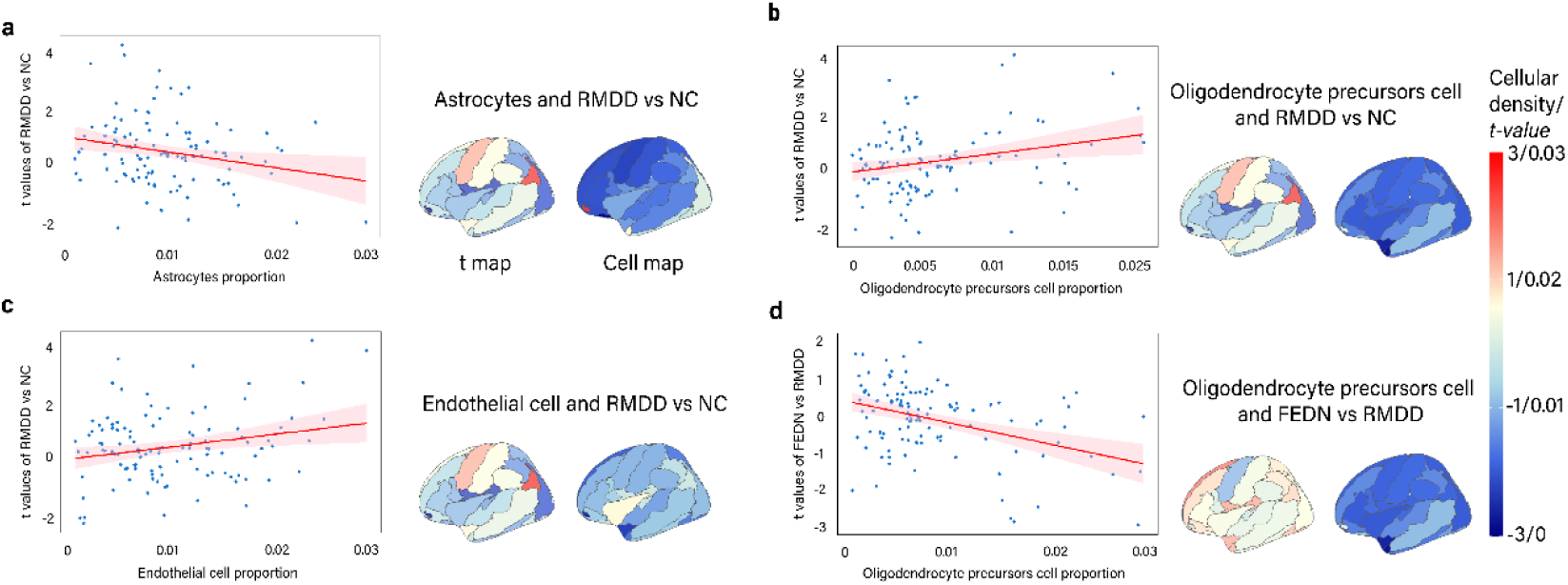
Spatial associations between FCS alterations and cell type proportions. (a-c) Astrocytes, endothelial cells, and OPCs showed significant negative correlations with FCS changes in RMDD vs. NC. (d) OPCs exhibited a significant positive correlation with FCS changes in FEDN vs. RMDD.

### 3.4 Correlation between MDD-related FCS alterations and neurotransmitters

Cross-regional spatial correlation analyses revealed distinct associations between FCS differences and neurotransmitter density distributions in MDD and its subtypes (Fig. 4a). The FCS alterations in total MDD exhibited significant negative correlations with serotonin 5-hydroxytryptamine receptor subtype 4 (5-HT4; r = −0.20, adjusted-p = 0.028), dopamine transporter (DAT; r = −0.24, adjusted-p = 0.007), dopamine synthesis capacity (F-DOPA; r = −0.25, adjusted-p = 0.005), and gamma-aminobutyric acid (GABA-a; r = −0.19, adjusted-p = 0.041). When dividing MDD into subgroups, significant negative correlations were observed for the RMDD vs. NC comparison with 5-HT1a (r = −0.40, adjusted-p = 0.002), 5-HT2a (r = −0.26, adjusted-p = 0.04), GABA-a (r = −0.24, adjusted-p = 0.01), and kappa opioid receptor (KappaOp; r = −0.38, adjusted-p = 0.002). For FEDN vs. RMDD comparison, significant correlations were identified with 5-HT1a (r = 0.40, adjusted-p = 0.002), 5-HT2a (r = 0.38, adjusted-p = 0.04), GABA-a (r = 0.20, adjusted-p = 0.04), KappaOp (r = 0.27, adjusted-p = 0.04), metabotropic glutamate receptor subtype 5 (mGluR5; r = 0.30, adjusted-p = 0.03), DAT (r = −0.19, adjusted-p = 0.03), norepinephrine transporter (NAT; r = −0.20, adjusted-p = 0.03), serotonin transporter (SERT; r = −0.24, adjusted-p = 0.009), and vesicular acetylcholine transporter (VAChT; r = −0.22, adjusted-p = 0.02). However, no significant correlations were detected in the FEDN vs. NC comparison. The density distributions of 5-HT1a, 5-HT2a, GABA-a, and KappaOp, which exhibited the most correlations with FCS differences among MDD subtypes, are presented in Fig. 4b.

### 3.5 Cell Types Associated with FCS Alterations in MDD Subtypes

Whole-brain distribution maps for six distinct cell types are illustrated in Supplementary Fig. 11. FCS differences between RMDD and NC exhibited significant correlations with the distribution of various cell types, including astrocytes (r = −0.23, adjusted-p = 0.03), endothelial cells (r = 0.23, adjusted-p = 0.04), and oligodendrocyte precursor cells (OPCs) (r = 0.26, adjusted-p = 0.04) (Fig. 5a-c). Conversely, the FCS difference between FEDN and RMDD demonstrated a significant negative correlation with the distribution of OPCs (r = −0.40, adjusted-p = 6.18×10^-5^) (Fig. 5d). No significant correlations were observed in the distribution of cell types with FCS differences among other groups.

### 3.6 Validation Results

The validation cohort revealed increased FCS in the bilateral hippocampus, parahippocampal gyrus, and right superior temporal pole, and decreased FCS in the right postcentral gyrus, bilateral anterior cingulate cortex, left paracentral lobule, and left angular gyrus in MDD compared to NCs, without accounting for distance effects. In terms of distance-dependent effects, we identified increased short-range FCS in the left angular gyrus, right middle temporal pole, bilateral precentral gyrus, fusiform gyrus, parahippocampal gyrus, and medial temporal lobe, as well as decreased FCS in the left calcarine fissure in MDD relative to NCs. Additionally, increases in the mid-cingulum and parahippocampal regions, and decreases in the primary motor cortex were observed for medium-range FCS. These findings are partially consistent with the primary findings from the comparison between total MDD and NC in the discovery cohort (Supplementary Fig. 2).

The regional gene expression profiles and their correlations with FCS alterations in MDD vs. NC were replicated in the validation cohort. The gene enrichment analysis was highly similar to those observed in the RMDD vs. NC and FEDN vs. RMDD comparisons in the discovery cohort (PLS1 for RMDD vs. NC and PLS2 for FEDN vs. RMDD), further demonstrating the biological processes related to chemical synaptic transmission and the regulation of transcription by RNA polymerase II, as well as the molecular functions of DNA binding underlying FCS differences between MDD subtypes (Supplementary Fig. 8 and 9).

The validation of neurotransmitter-related results confirmed the findings of MDD vs. NC, RMDD vs. NC, and FEDN vs. RMDD comparisons, demonstrating significant negative correlations between FCS alterations and 5-HT2a (r = −0.34, adjusted-p = 0.03), GABA-a (r = −0.23, adjusted-p = 0.02), NAT (r = −0.30, adjusted-p = 0.002), and NMDA (r = −0.19, adjusted-p = 0.04) (Supplementary Fig. 10). These results suggest a robust association between disruptions of FC in MDD and dysregulation of neurotransmitter systems. Regarding cell type-specific findings, the validation cohort identified a significant positive correlation between FCS differences and the distributions of neurons (r = 0.27, adjusted-p = 0.02, Supplementary Fig. 12).

## 4. DISCUSSION

In this study, we investigated the multi-scale pathophysiological mechanisms underlying major depressive disorder (MDD) subtypes by integrating functional connectivity, transcriptomic, neurotransmitter, and cell-type analyses across two large Asian cohorts. Our findings revealed distinct patterns of distance-dependent FCS alterations in MDD and RMDD. Furthermore, we found that the FCS differences between FEDN and RMDD were associated with the neuron projection and synaptic signaling, the monoaminergic and GABAergic systems, and the astrocytes, and endothelial cells. These results provide novel insights into the neurobiological underpinnings of MDD subtypes and highlight potential avenues for targeted interventions.

### 4.1 Distance-dependent FCS Alterations: A Novel Perspective on MDD Dysconnection

Without considering distance effect, we observed increased FCS in the left angular gyrus for total MDD, and in the right posterior cingulate gyrus, bilateral thalamus, and cerebellum for RMDD compared to NC. In contrast, this study uncovered complex patterns of dysconnectivity across different spatial scales. Specifically, we found short- and medium-range hyperconnectivity within the default mode network (DMN), salience network, and cognitive control network in total MDD patients. These findings extend recent studies reporting decreased FC within the DMN ^4^, while highlighting increased short-range FC in frontal and parietal regions ^59^. When classifying MDD into subtypes, RMDD patients exhibited extensive long-range hyperconnectivity, whereas FEDN patients did not show significant distance-dependent changes. This aligns with findings of functional alterations in both reward and default mode networks in RMDD ^2,60–62^, while FEDN showed no significant changes compared to NC^2,60,62^. Moreover, the hyperconnectivity across different distance ranges in RMDD may reflect a shift toward a more random network organization, consistent with studies reporting a disruption in the balance between local specialization and global integration in MDD patients ^63–65^. The lack of significant differences between FEDN and medicated first-episode patients in our study suggests that the observed alterations in RMDD are likely due to disease progression rather than medication effects, aligning with observations from longitudinal studies ^60,66,67^.

Our findings support the conclusion that functional network alterations in MDD evolve spatially and temporally. Studies examining the progression of network disruptions in MDD have found that alterations typically begin in the subgenual anterior cingulate cortex and gradually spread to wider networks across the brain ^64,65,68^. This observation aligns with recent research identifying distinct spatiotemporal MDD subtypes originating from the subgenual anterior cingulate cortex, hippocampus, and superior frontal gyrus ^69^. The extensive long-range hyperconnectivity in RMDD patients corresponds well with both anterior cingulate cortex-led and frontal-led subtypes, suggesting common pathways in the development of certain MDD subtypes. Additionally, our findings underscore that the progression of MDD and its subtypes involves complex changes in brain networks across different spatial scales. Collectively, these distinct patterns of dysconnectivity across MDD subtypes provide new insights into the neurobiological basis of MDD progression.

### 4.2 Transcriptomic Signatures: Linking Gene Expression to Dysconnection in MDD

Our transcriptomic analysis revealed that genes associated with FCS differences between FEDN and RMDD were enriched for biological processes related to chemical synaptic transmission and plasma membrane bounded neuron projection. This finding supports that synaptic dysfunction is a core feature across depressive disorders ^9^ and that plasma membrane-bound cell projections are related to neuroplasticity changes in MDD ^70,71^. The genes related to FCS differences between RMDD and NC uniquely enriched for RNA polymerase II-related transcription regulation, aligning with studies implicating epigenetic mechanisms ^72^, while those associated with FCS differences between FEDN and NC were specifically enriched in biological processes and cellular components related to mitochondrial function, consistent with previous studies reporting reduced mitochondrial respiratory chain function in depression ^73–76^. In summary, these findings indicate that distinct molecular basis may underlie the macroscale FCS differences between MDD subtypes.

Interestingly, we identified several genes most associated with the FCS difference between RMDD and FEDN, including DPF3, CECR2, USP3, and CHRM3. DPF3, involved in chromatin remodeling ^77^, has been implicated in stress-induced depression-like behaviors ^78^. Several studies have demonstrated that DPF3 regulates the expression of genes involved in synaptic plasticity and neurotransmission ^79–81^, providing a potential link between epigenetic mechanisms and the observed FCS alterations in our study. CECR2, another chromatin remodeling factor ^82^, and USP3, a deubiquitinating enzyme involved in cellular stress responses ^83^, further support the involvement of epigenetic mechanisms in MDD progression ^72^. The identification of CHRM3, which encodes the M3 muscarinic acetylcholine receptor ^84^, is particularly relevant given recent evidence of cholinergic system involvement in rapid antidepressant responses^85^. Research indicates that activation of muscarinic acetylcholine receptors in the hippocampus mediates ketamine’s rapid antidepressant effects ^86^, suggesting a potential target for subtype-specific interventions based on gene expression patterns we observed.

### 4.3 Neurotransmitter and Cellular Correlates: Integrating Multiple Levels of Analysis

The neurotransmitter correlation analysis provided evidence for the involvement of multiple neurotransmitter systems in MDD pathophysiology. We observed negative correlations between GABAergic as well as KappaOp receptors densities and FCS differences in RMDD vs. NC but not in FEDN vs. NC and FEM vs. FEDN, consistent with recent studies on neurotransmitter imbalances in MDD ^87–89^, suggesting these two receptors may be associated with MDD severity or duration of illness rather than drug effect. Interestingly, we found extensive overlap of neurotransmitters related to FCS differences in FEDN vs. RMDD and RMDD vs. NC, suggesting potential compensatory mechanisms among different MDD subtypes. These findings align with the growing recognition of the complex interplay between neurotransmitter systems in MDD ^90^ and support recent interest in multi-target pharmacological approaches ^91^. Recent research has proposed that rapid antidepressant effects involve modulation of multiple neurotransmitter systems, including glutamate, GABA, and monoamines ^92,93^. We observed significant correlations between FCS differences and multiple neurotransmitter systems in FEM vs. FEDN, including serotonergic, dopaminergic, glutamatergic, and cholinergic systems. This multi-system pattern aligns with evidence that selective serotonin reuptake inhibitors and other antidepressants not only regulate their primary target systems but also induce cascading effects on other neurotransmitter pathways, as clinical studies have demonstrated that escitalopram treatment simultaneously modulates serotonergic and dopaminergic transmission, while long-term antidepressant treatment affects glutamatergic neurotransmission through synaptic plasticity mechanisms^94–96^. Our results provide a neuroimaging correlate to these molecular findings, suggesting that the observed FCS alterations may reflect the cumulative effects of neurotransmitter imbalances across MDD subtypes.

Our cell-type-specific analysis revealed associations between the distributions of astrocytes, endothelial cells, and oligodendrocyte precursor cells (OPCs) and FCS differences in RMDD compared to NC, but not in comparisons involving FEDN. These findings support the emerging concept of MDD as a disorder involving multiple cell types beyond neurons ^28,97^. The involvement of astrocytes aligns with recent evidence of their role in synaptic plasticity and neurotransmitter metabolism in MDD ^98^. Studies have demonstrated that astrocyte-specific alterations can lead to depressive-like behaviors in animal models ^99,100^, providing a potential mechanism linking astrocyte dysfunction to the FCS alterations observed we. The association with endothelial cells supports the vascular depression hypothesis ^101^, while findings related to OPCs suggest potential white matter changes in MDD progression ^102^. The interplay between neurotransmitter systems and cell-type distributions may significantly influence the observed FCS alterations. For instance, the negative correlation between FCS alterations and GABAergic receptor density, coupled with the involvement of astrocytes, suggests a mechanism where astrocyte dysfunction leads to altered GABA signaling, which in turn affects depressive symptoms ^103^. Furthermore, while OPCs showed a significant correlation with FCS differences in RMDD compared to FEDN, they also correlated with those in FEM) compared to FEDN, indicating that this cell type may be more associated with FCS alterations induced by drug treatment rather than depressive severity. Collectively, these cellular and neurotransmitter correlates provide a biological basis for the observed FCS alterations and highlight the complex, multi-level nature of MDD pathophysiology.

### 4.4 Cross-Cultural Considerations

Given the known impact of cultural factors on the presentation and perception of depressive symptoms ^104,105^, our study utilized two large Asian cohorts to observe several MDD-related dysconnections that differ from those reported in studies involving Western populations ^42–46^. For instance, we found prominent short- and medium-range hyperconnectivity in MDD, with additional long-range hyperconnectivity specifically in RMDD, whereas studies based on Western populations reported mixed patterns of hypo- and hyperconnectivity ^106,107^. Supporting evidence from our transcriptomic findings, we revealed enrichment in synaptic transmission-related genes, similar to findings in Western studies ^108^, while we identified a unique enrichment pattern related to RNA polymerase II-mediated transcription in RMDD. This observation aligns with recent genetic studies that highlight distinct risk loci for MDD in European and Asian populations ^109^. A large-scale genetic study identified two depression risk loci in Han Chinese women that were not previously reported in European populations, underscoring the importance of population-specific genetic studies in understanding MDD^110^.

The validation of our key findings across two independent Asian cohorts strengthens the reliability of our results and suggests their generalizability across East Asian populations. Notably, the replication of PLS1 gene enrichment results for both RMDD vs. NC and FEDN vs. RMDD comparisons in an independent Japanese cohort provides robust support for the validity of our transcriptomic findings.

### 4.5 Limitations and Future Directions

Despite the strengths of our study, several limitations should be acknowledged:

1. The cross-sectional nature of our data limits causal inferences. Longitudinal studies tracking individuals from FEDN to RMDD stages could provide valuable insights into the temporal dynamics of the observed FCS alterations.
2. A significant limitation is that the neuroimaging, transcriptomic, and neurotransmitter data used in this study were derived from different sources and individuals. This non-matched approach may introduce potential biases and limit the interpretation of relationships between these different modalities.
3. The use of data from the AHBA, derived from postmortem samples without MDD diagnoses, limits our ability to examine transcriptome-neuroimaging associations across subjects. However, comparative studies have shown a high degree of conservation in overall gene expression across human populations ^111^, suggesting that the identified FCS-related genes are likely conserved across subjects.
4. Our validation cohort did not include MDD subtypes, which limits our ability to confirm subtype-specific findings in an independent sample.
5. Our study focused on Asian populations, which may limit the generalizability of our findings to other ethnic groups.
6. While our multi-modal approach provides a comprehensive view of MDD, incorporating additional data types, such as structural connectivity or task-based fMRI, could yield richer insights into the disorder.

Future studies should aim to address these limitations by:

1. Employing matched multi-modal data from the same individuals, particularly those with MDD, to enhance insights into the relationships between brain structure, function, and molecular mechanisms in MDD.
2. Including well-characterized MDD subtypes in both discovery and validation cohorts to improve the reliability and specificity of subtype-related findings.
3. Directly comparing Asian and Western cohorts to elucidate population-specific aspects of MDD pathophysiology.
4. Incorporating additional neuroimaging modalities to deepen our understanding of the complex interplay between brain structure, function, and molecular mechanisms in MDD.

## 5. CONCLUSIONS

In conclusion, our study provides a multi-scale characterization of functional dysconnections in MDD across two Asian cohorts. We revealed distinct patterns of distance-dependent FCS alterations associated with specific transcriptomic signatures, neurotransmitter systems, and cell types. These findings connect macroscale connectional alterations with microscale molecular and cellular features, offering valuable insights into the pathophysiological mechanisms of MDD subtypes in Asian populations.

## Acknowledgments

This work is supported by the Chu Kochen Honors College Foundation.

## Data and Code Availability

Data of the REST-meta-MDD project are available at : http://rfmri.org/REST-meta-MDD. Data of the validation cohort are available at Decoded Neurofeedback (DecNef) Project Brain Data Repository (https://bicr.atr.jp/decnefpro/data). Neurotransmitter receptor and transporter data can be obtained online at https://github.com/juryxy/JuSpace/tree/JuSpace_v1.5/JuSpace_v1.5/PETatlas. Human gene expression data that support the findings of this study are available in the Allen Brain Atlas (https://human.brain-map.org/static/download).

## MRI Data of Both Cohorts

The MRI data of discovery cohort was from REST-meta-MDD Project, see the detail information of data acquisition, subjects selection and preprocessing at https://doi.org/10.57760/sciencedb.o00115.00013 The MRI data of validation cohort was from Decoded Neurofeedback (DecNef) Project. The data form is a collection of inter-regional correlation matrices based on the resting-state functional MRI (rsfMRI) data. We only selected the data acquired at the University of Tokyo Hospital between the years 2014 and 2017, due to this site provides matrix calculated using Automated Anatomical Labeling (AAL) Atlas, which has the lowest transcription loss rate.

## MRI Data Acquisition

To find details about the discovery cohort in the REST-meta-MDD project, including consortium sites, contributors, sample size, data acquisition parameters, and published studies, please refer to Supplementary Table S3 of the paper titled *“Reduced default mode network functional connectivity in patients with recurrent major depressive disorder.”* This table should provide comprehensive information on the dataset and the methods used in this study.

For validation cohort GE MR750W 3.0T system at the University of Tokyo Hospital Protocol. The scan parameters for the rs-fMRI images were as follows: TR = 2500 ms; TE = 30 ms; flip angle = 80°; matrix = 64 x 64; FOV = 212 mm × 212 mm; in-place resolution = 3.3 mm × 3.3 mm; slice thickness = 3.2 mm; gap = 0.8 mm; number of slices = 40; number of volumes = 240; scan time = 10 mins.

**Supplementary Table 1.** Demographic characteristics of validation cohort.

| Grand Analysis of Age |  |  |  |  |  |
| --- | --- | --- | --- | --- | --- |
| MDD (N=62) |  | NC (N=170) |  | Mann-Whitney |  |
| min | 18 | min | 19 | P | <0.001 |
| max | 63 | max | 80 | Z | 2.282 |
| mean | 38.74 | mean | 35.59 | U | 8495.5 |
| SD | 11.62 | SD | 17.5 |  |  |
| Grand Analysis of Sex |  |  |  |  |  |
| MDD (N=62) |  |  | NC (N=170) |  |  |
| Male | 36 | 58.06% | Male | 78 | 45.88% |
| Female | 26 | 41.94% | Female | 92 | 54.12% |
| Chi-Square |  |  |  |  |  |
| | $\chi^2$ | | | | 2.2322 |
|  | P |  |  |  | 0.1351 |

**Supplementary Fig.1.**
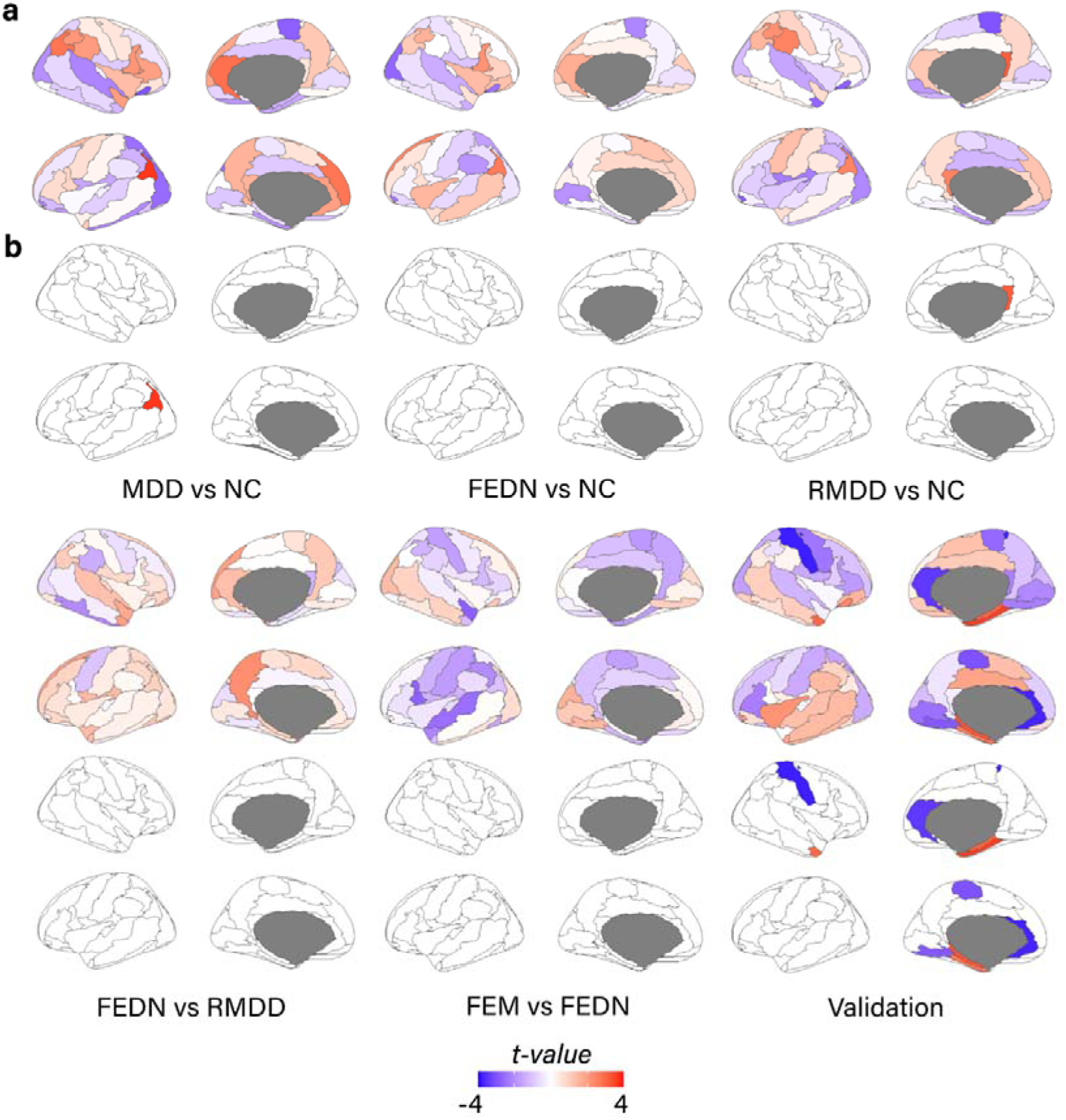
The FCS alterations without distance effects. **a** No-distance effects t-values of MDD vs. NC (848 MDDs and 794 NCs) comparison, FEDN vs. NC (232 FEDNs and 394 NCs), RMDD vs. NC (189 RMDDs and 427 NCs), FEDN vs. RMDD (FEDNs and 72 RMDDs), FEM vs. FEDN (100 FEMs and 227 FEDNs) and validation cohort (62 MDDs and 170 NCs). **b** Brain regions that demonstrated statistically significant changes after adjusting for multiple comparisons using FDR correction.

**Supplementary Fig.2.**
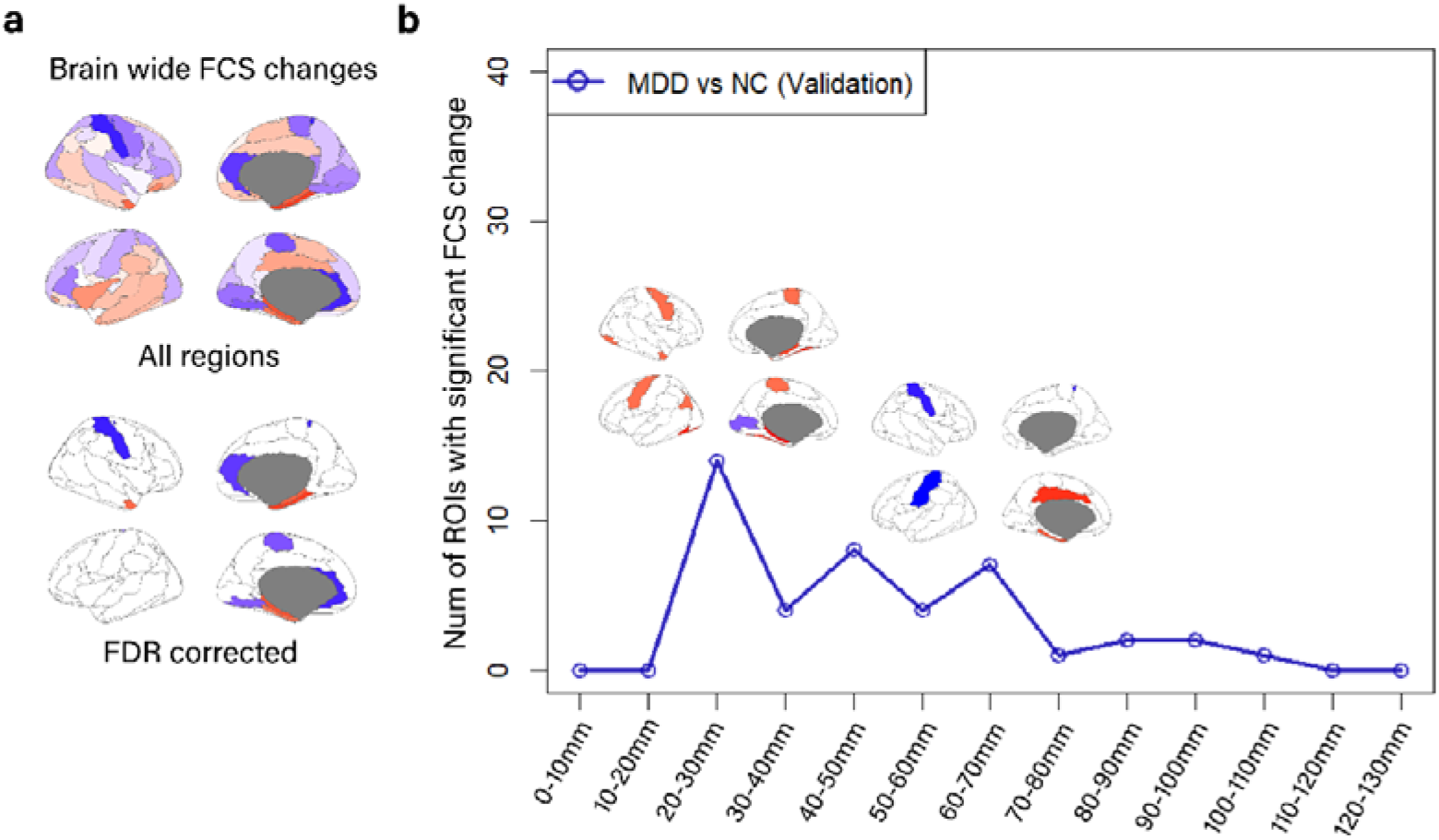
Brain-wide and distance-dependent FCS changes in the MDD from validation cohorts. **a** Brain-wide FCS alterations without considering distance dependency. Upper row displays all regions with FCS changes, while bottom row shows regions surviving FDR correction. **b** Distance-dependent FCS alterations, which showed obvious short and medium heterogeneity in MDD vs. NC (Validation). The x-axis represents the Euclidean distance between brain regions ranging from 0 to 130mm. The y-axis indicates the number of ROIs with significant FCS changes. For all brain images, the colorbar represents t values, and positive and negative values indicate higher and lower FCS in former group than latter group, respectively.

**Supplementary Fig.3.**
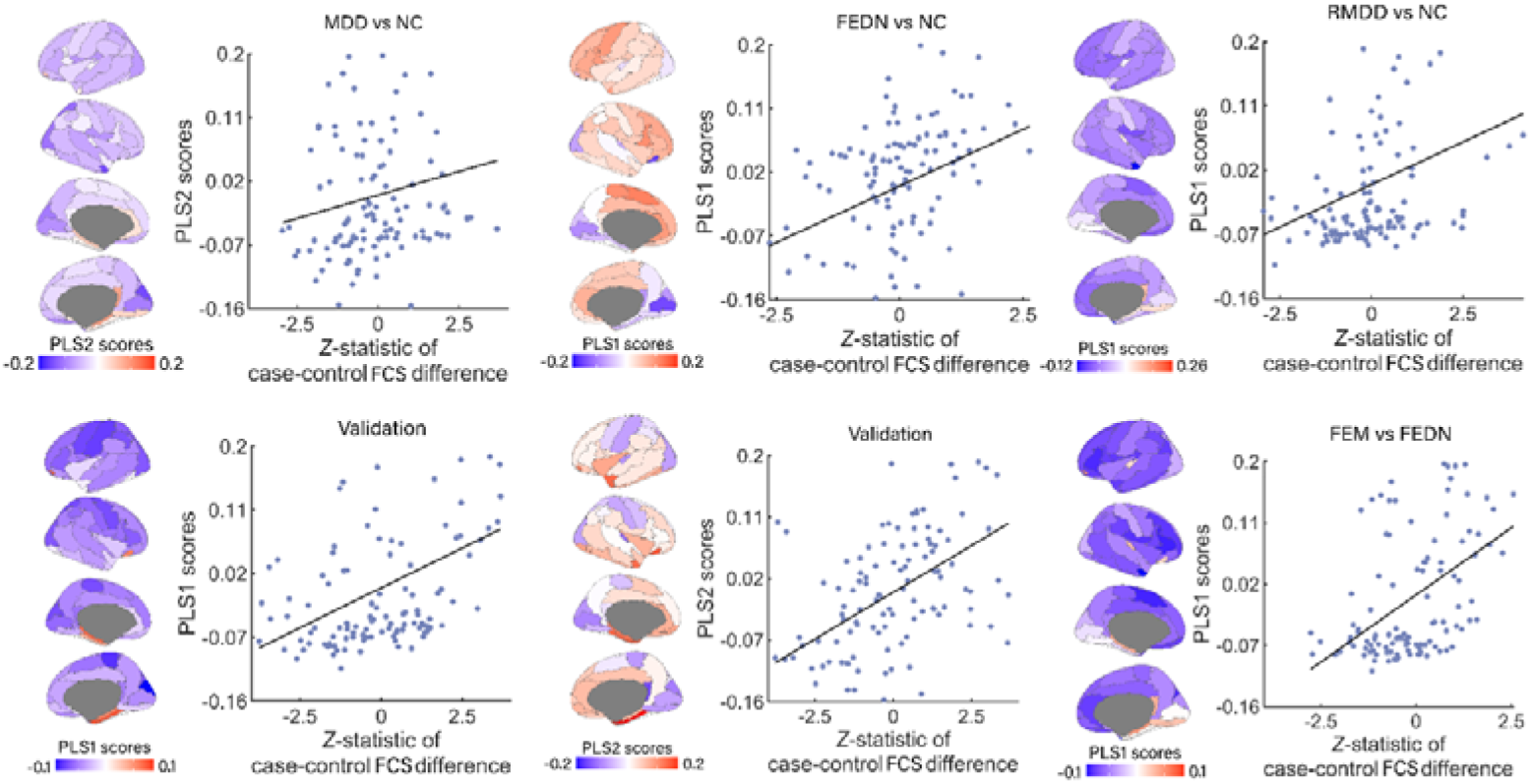
The expression profile of each PLS component in discovery and validation groups. The transcriptional profiles were positively correlated with the between-group t values of FCS differences. The PLS scores shows gene expression level across the cortex.

**Supplementary Fig.4.**
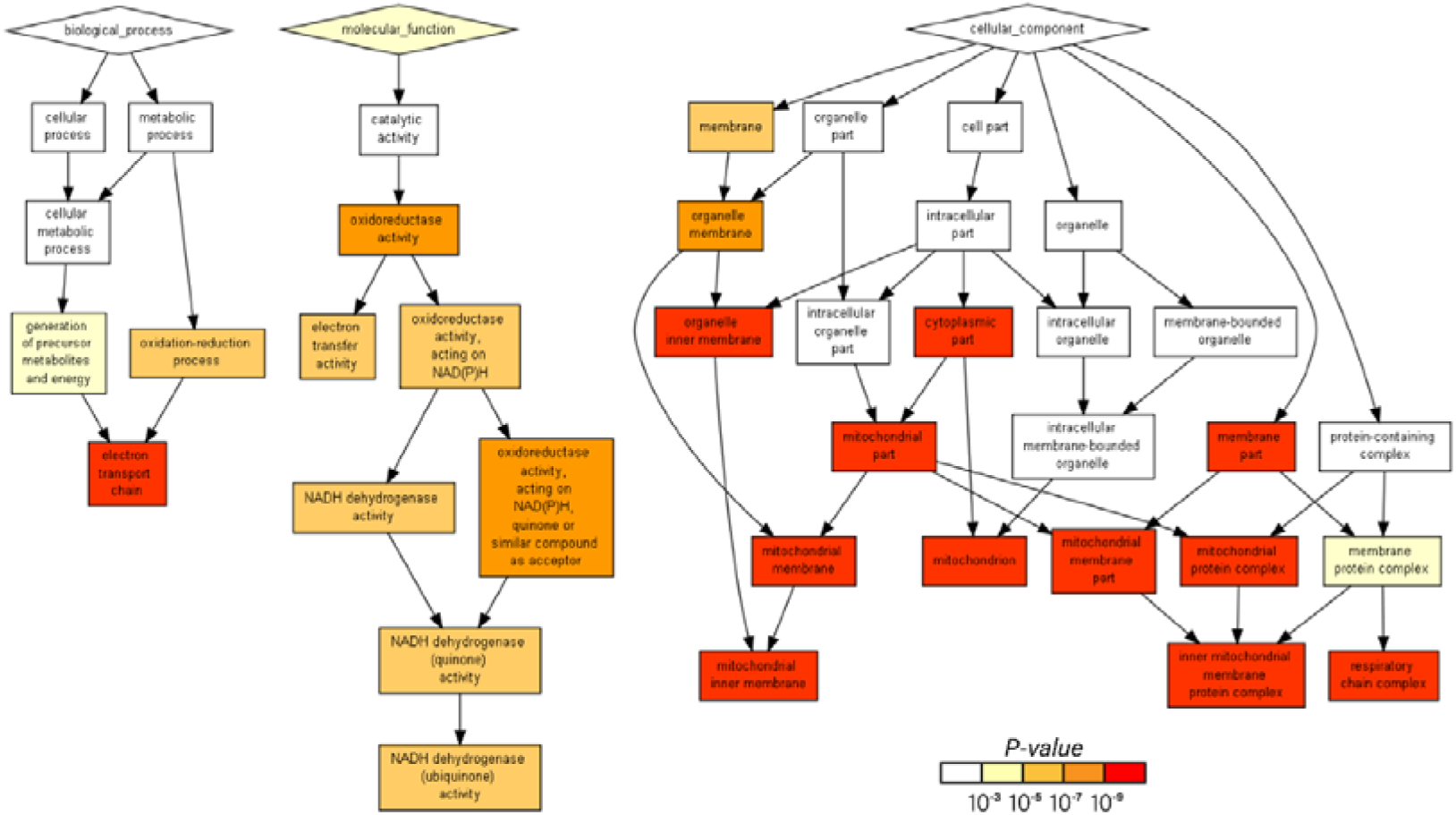
PLS1 gene enrichment analysis results in FEDN vs. NC. PLS1 genes of FEDN vs. NC were enriched in biological processes of electron transport chain, molecular function of NADH dehydrogenase activity, and cell component of mitochondrial inner membrane protein complex and respiratory chain complex.

**Supplementary Fig.5.**
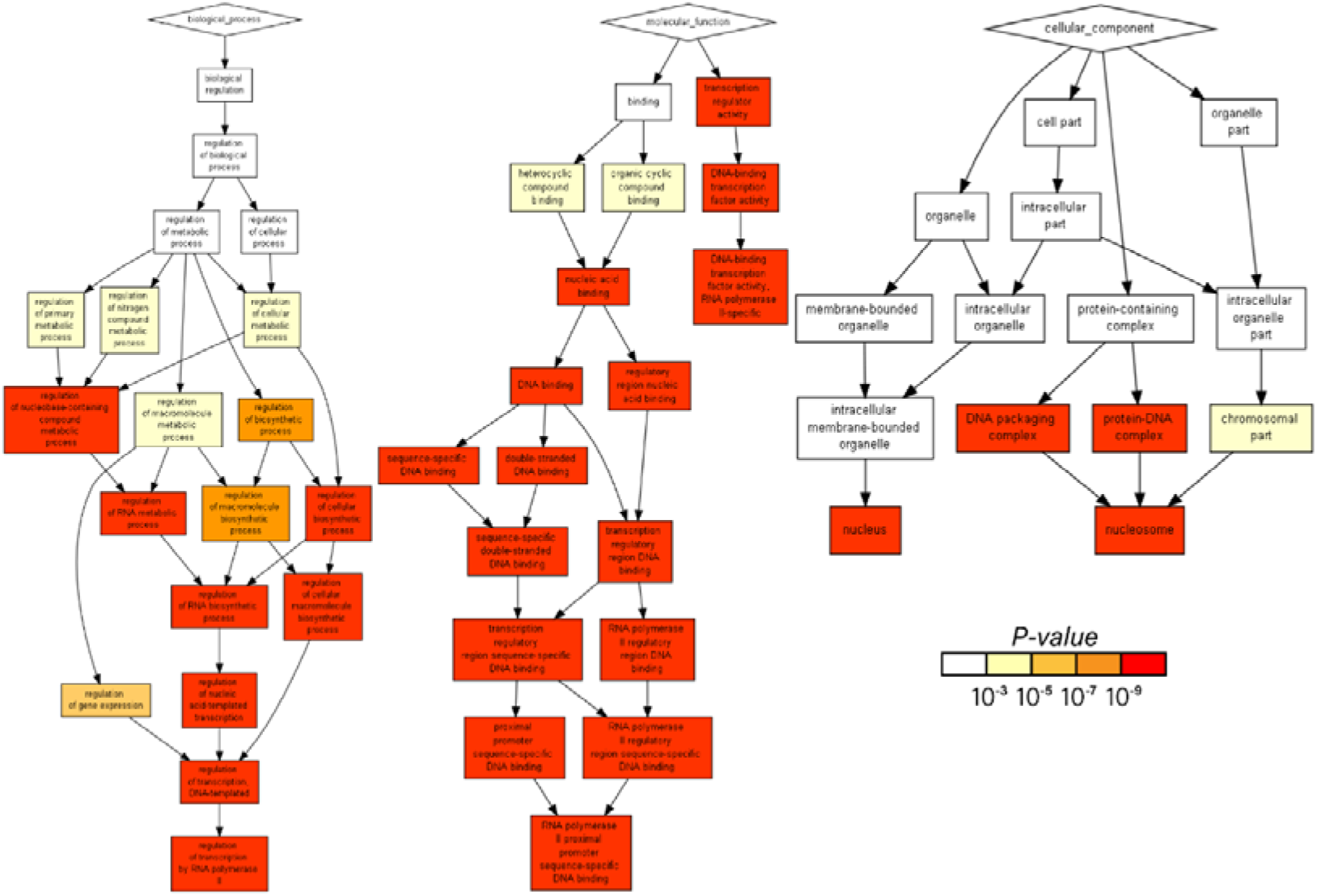
PLS1 gene enrichment analysis results in RMDD vs. NC. PLS1 genes of RMDD vs. NC were enriched in biological processes related to chemical synaptic transmission, molecular function of RNA polymerase Il proximal promoter sequence-specific DNA binding and cell component of nucleus.

**Supplementary Fig.6.**
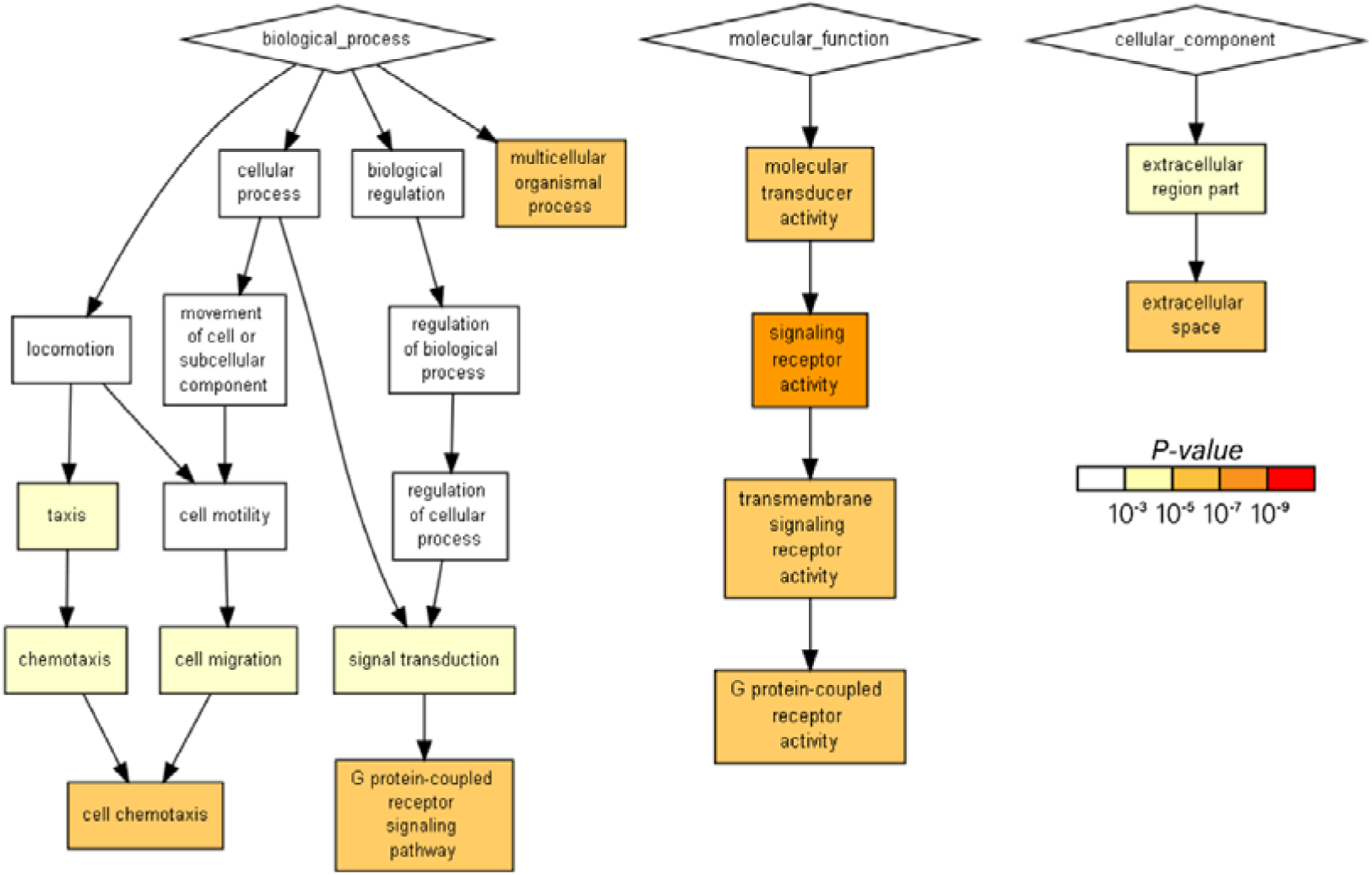
PLS2 gene enrichment analysis results in MDD vs. NC. PLS2 genes of MDD vs. NC were enriched in biological processes of cell chemotaxis, G protein-coupled receptor signaling pathway and multicellular organismal process, molecular function of G protein-coupled receptor activity, and cell component of extracellular space.

**Supplementary Fig.7.**
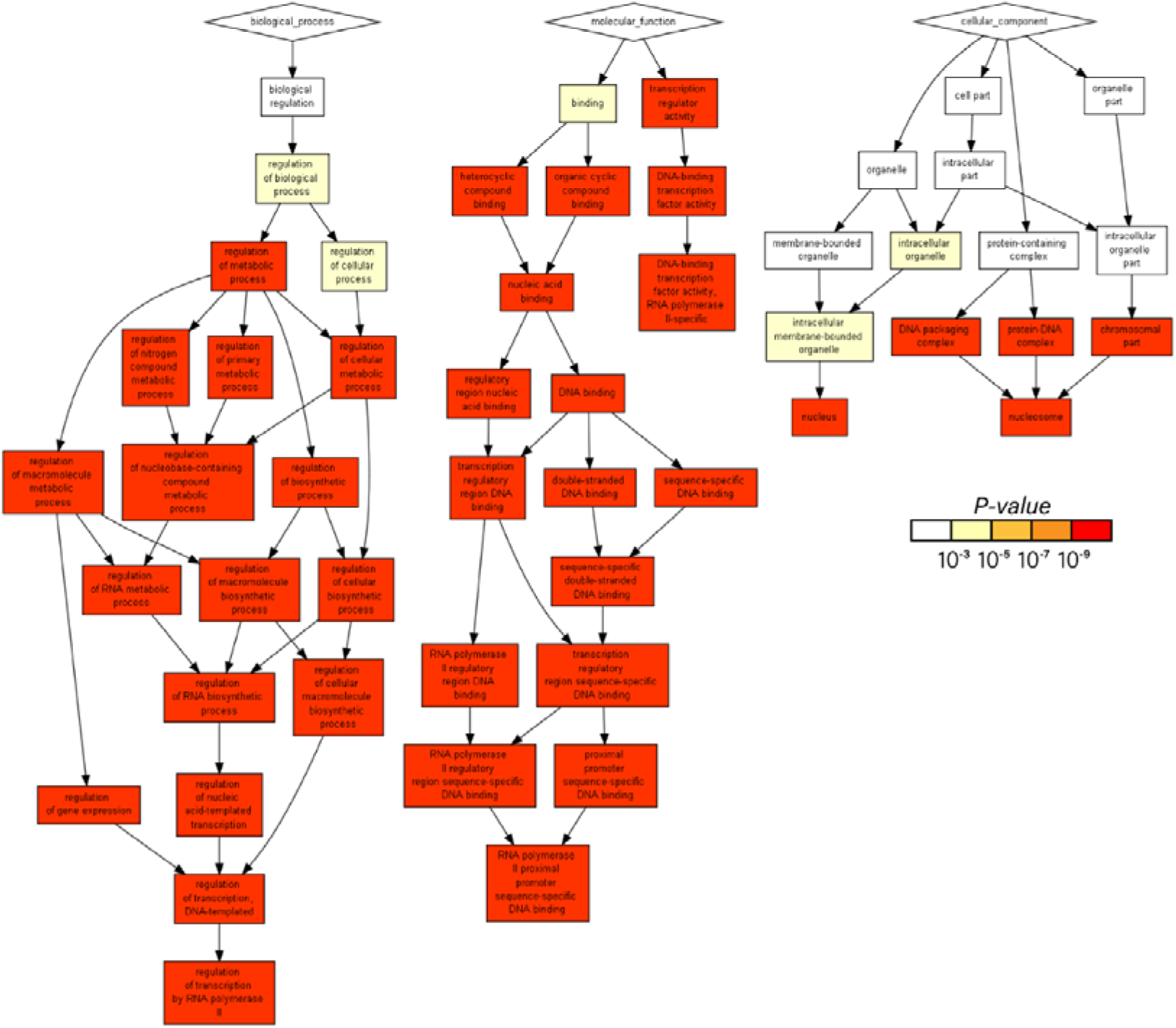
PLS1 gene enrichment analysis results in FEM vs FEDN. PLS1 genes of validation cohort were enriched in biological processes of regulation of transcription by RNA polymerase ll, molecular function of RNA polymerase ll proximal promoter sequence-specific DNA binding, and cell component of nucleus and nucleosome.

**Supplementary Fig.8.**
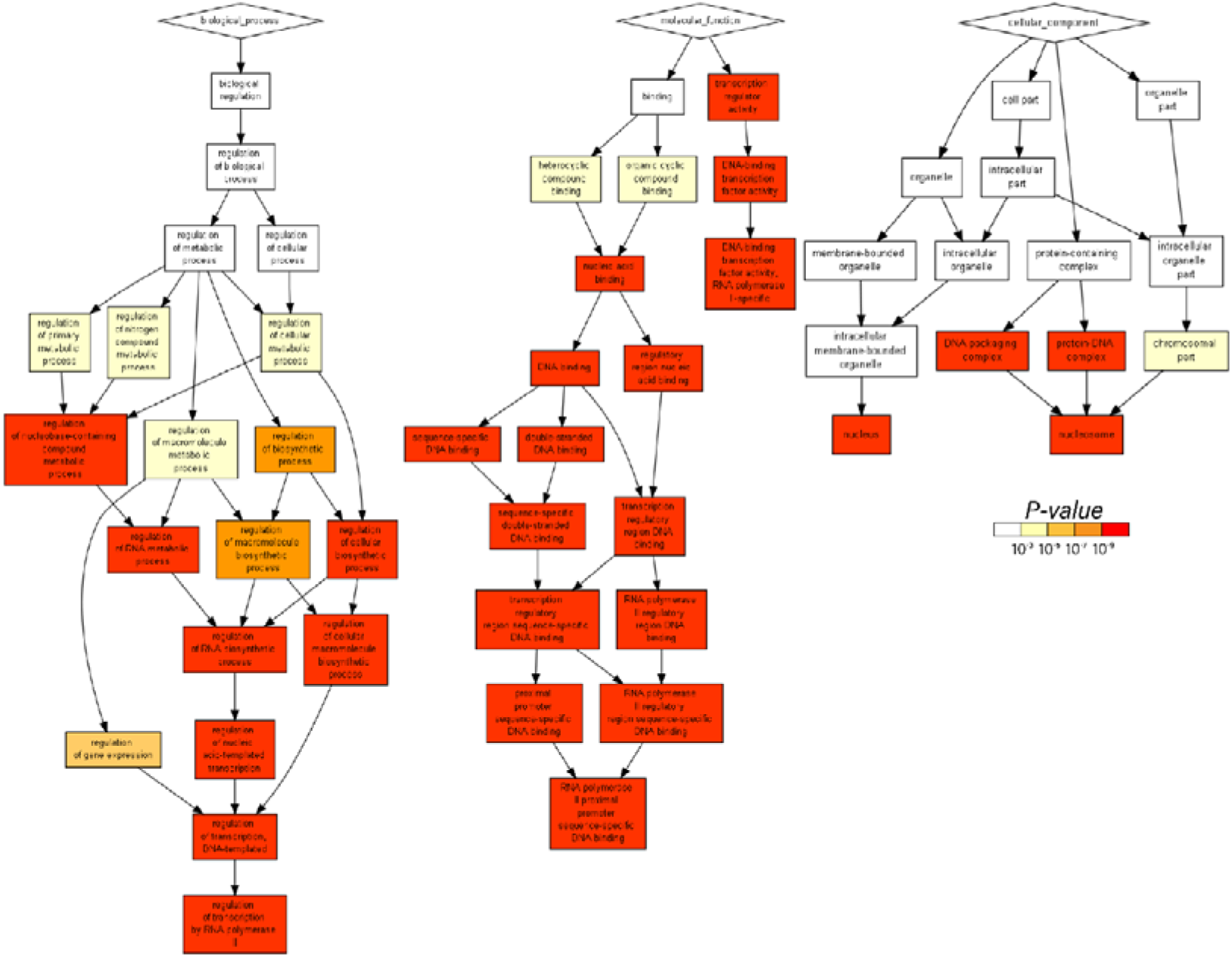
PLS1 gene enrichment analysis results in validation cohort. PLS1 genes of validation cohort were enriched in biological processes of regulation of transcription by RNA polymerase ll, molecular function of RNA polymerase ll proximal promoter sequence-specifc DNA binding, and cell component of nucleus and nucleosome.

**Supplementary Fig.9.**
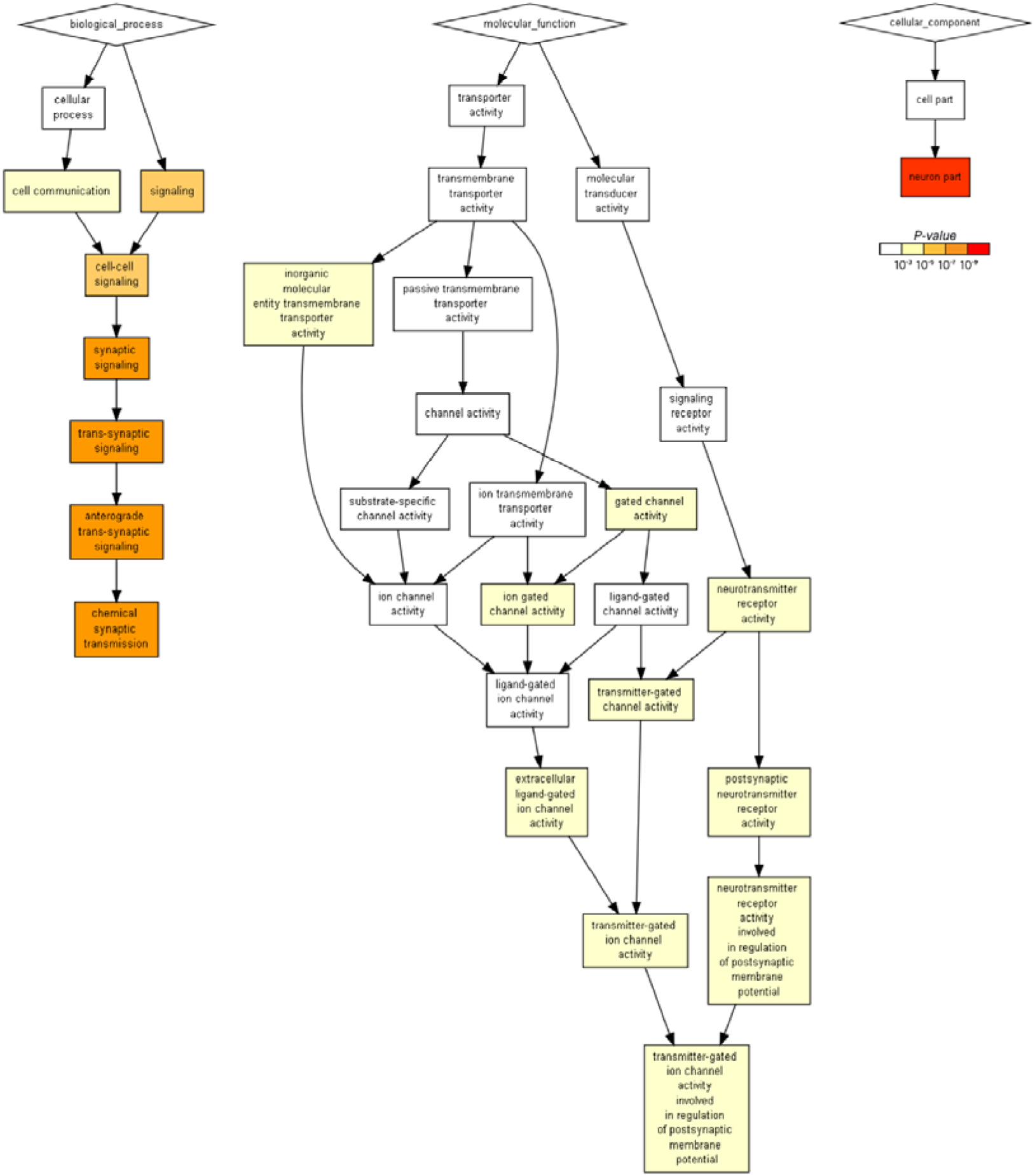
PLS2 gene enrichment analysis results in validation cohort. PLS2 genes of validation cohort were enriched in biological processes of chemical synaptic transmission, molecular function of transmitter-gated ion channel activity involved in regulation of postsynaptic membrane potential, and cell component of neuron.

**Supplementary Fig.10.**
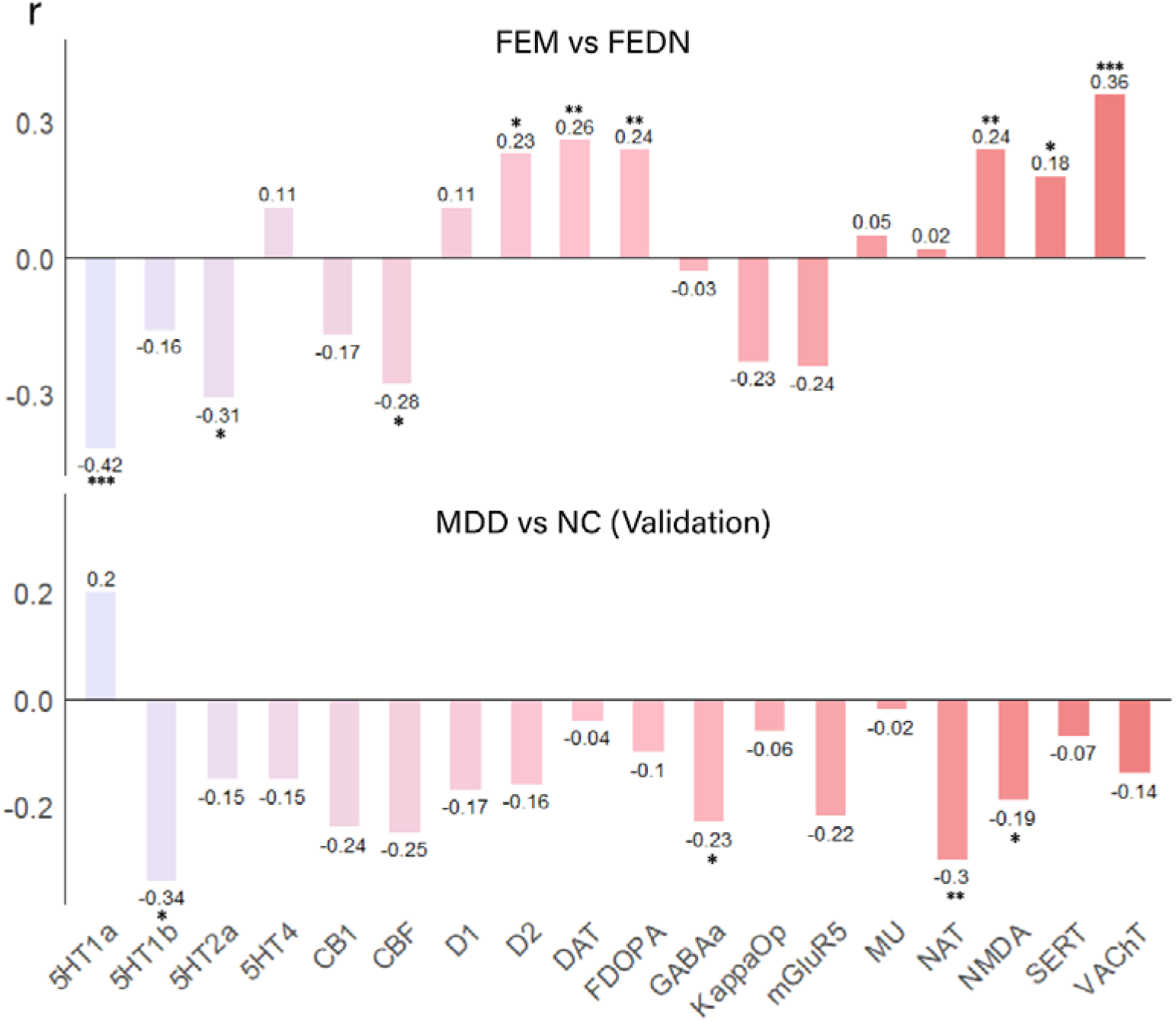
The neurotransmitter receptors/transporters correlation results in FEM vs. FEDN and validation cohort. For FEM vs. FEDN, Significant negative correlations were observed between FCS alterations and 5-HT2a (r = −0.42, p = 0.0001), 5-HT2a (r = −0.31, p = 0.02), CBF (r = −0.28, p = 0.03); while significant positive correlations with D2 (r = 0.23, p = 0.02), DAT (r = 0.26, p = 0.003), FDOPA (r = 0.24, p = 0.008), NMDA (r = 0.24, p = 0.008), SERT (r = 0.18, p = 0.04) and VAChT (r = 0.36, p = 0.0001). For validation cohort, Significant negative correlations were observed between FCS alterations and 5-HT2a (r = −0.34, p = 0.03), GABA-a (r = −0.23, p = 0.02), NAT (r = −0.30, p = 0.002), and NMDA (r = −0.19, p = 0.04).

**Supplementary Fig.11.**
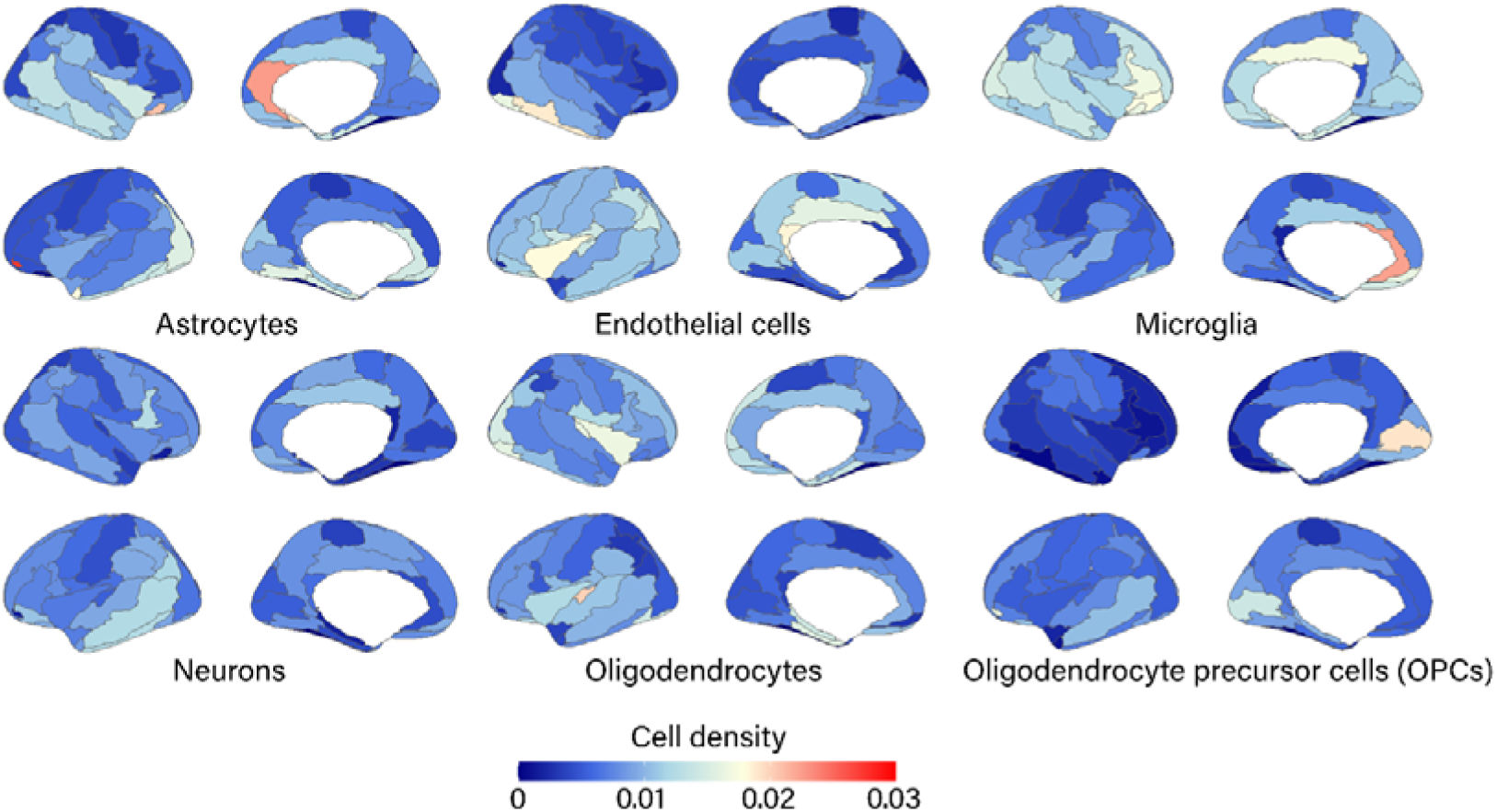
Whole-brain distribution maps for six distinct cell types. The deconvolution method is used to extract the proportions of six cell types from the transcriptome, including neurons, astrocytes, oligodendrocytes, microglia, endothelial cells, and oligodendrocyte precursor cells (OPCs). The 3D whole-brain maps of cellular abundance for these canonical cell types were sourced from the work of Veronika Pak et al. (https://github.com/neuropm-lab/cellmaps)

**Supplementary Fig.12.**
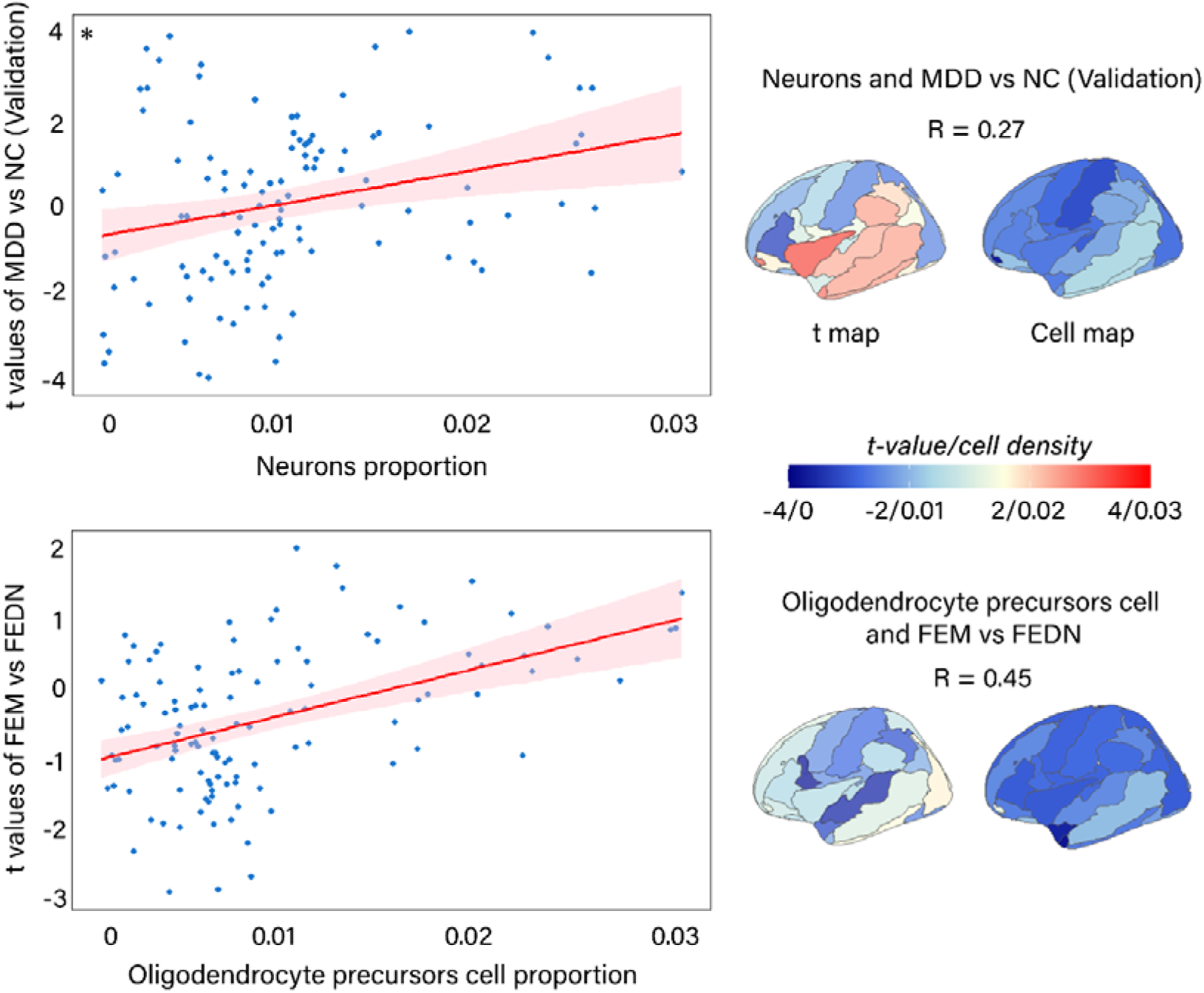
Spatial associations between FCS alterations and cell types proportion in FEM vs. FEDN and validation cohort. For FEM vs. FEDN, significant positive correlation showed between FCS changes and the proportion of OPCs (*r* = 0.45, *p* = 2.6e-7). For validation cohort, significant positive correlation showed between FCS changes and the proportion of neurons (*r* = 0.27, *p* = 0.0195).

